# Convergent abnormalities of β-amyloid deposition, glucose metabolism, and fMRI activity in the dorsal precuneus in subjective cognitive decline

**DOI:** 10.1101/2021.04.14.21255317

**Authors:** Xuan-Yu Li, Li-Xia Yuan, Fei-Fan Zhou, Chang-Chang Ding, Teng-Fei Guo, Wen-Ying Du, Jie-Hui Jiang, Frank Jessen, Yu-Feng Zang, Ying Han

## Abstract

**Background:** There has been no report on convergent local abnormalities of multiple functional brain imaging modalities including β-amyloid (Aβ) deposition, glucose metabolism, and resting-state functional magnetic resonance imaging (RS-fMRI) activities for participants with subjective cognitive decline (SCD).

**Methods:** Fifty participants with SCD and 15 normal controls (NC) were scanned with both [^18^F]-florbetapir positron emission tomography (PET) and [^18^F]-fluorodeoxyglucose PET, each PET sacn accompanied with simultaneous RS-fMRI. Voxel-wise metrics were analyzed, including Aβ deposition, glucose metabolism, and three local metrics for RS-fMRI, i.e., amplitude of low frequency fluctuation (ALFF), regional homogeneity (ReHo), and degree centrality (DC).

**Results:** The SCD group showed increased Aβ deposition and increased glucose metabolism (*P* < 0.05, corrected), as well as decreased ALFF, ReHo, and DC (*P* < 0.05, uncorrected) in the same area of the left dorsal precuneus (dPCu). The dPCu showed negative resting state functional connectivity (RSFC) with the default mode network (DMN). Regarding global Aβ deposition positivity, the Aβ deposition in the dPCu showed a gradient change, i.e., SCD^+^ > SCD^-^ > NC^-^. Further, both SCD^+^ and SCD^-^ showed increased glucose metabolism and decreased RS-fMRI metrics in the dPCu.

**Conclusions:** The convergent abnormal activities in the dPCu of SCD indicate that the dPCu is an early vulnerable region. The anti-RSFC of the dPCu with DMN supports that the earliest symptoms might be more related to other cognitive functions (e.g., unfocused attention) than episodic memory. (Funded by the National Key Research and Development Program of China and others; ClinicalTrials.gov number, NCT03370744.)

## Introduction

Subjective cognitive decline (SCD) refers to self-perceived cognitive decline relative to a previous status in the absence of measurable objective cognitive impairment.^1,2^ SCD may be the first symptomatic manifestation of Alzheimer’s disease (AD) and indicate a higher risk for a future objective cognitive impairment.^1^ Functional neuroimaging techniques, including β-amyloid (Aβ) positron emission tomography (PET), tau protein PET, [^18^F]-fluorodeoxyglucose (FDG)-PET, and resting-state functional magnetic resonance imaging (RS-fMRI), have been used in SCD studies. However, as summarized in the Supplementary Appendix (SA) Table S1, there has been no consensus on where the abnormal activity is in SCD.

The current study used a PET/MR hybrid scanner and analyzed the local activities including Aβ deposition, glucose metabolism, and three RS-fMRI local metrics, i.e., amplitude of low frequency fluctuation (ALFF), regional homogeneity (ReHo), and degree centrality (DC) in SCD and normal control (NC).^3-5^ Previous coordinate-based meta-analytic studies have suggested that the posterior medial parietal area in the default mode network (DMN) regions including the posterior cingulate cortex (PCC) and its adjacent precuneus (PCu) (usually named PCC/PCu in literatures) consistently showed increased Aβ deposition, decreased glucose metabolism, decreased ALFF, and decreased ReHo in the mild cognitive impairment (MCI) or AD.^6-9^ We therefore hypothesized that the PCC/PCu in SCD would show convergent abnormal activities.

## Methods

### Participants

Participants were recruited through public advertisements and referrals from general physicians, memory clinics, and informants. All participants signed a written informed consent before the experiment. The recruitment and assessment of participants have been described in the protocol of the Sino Longitudinal Study on Cognitive Decline (SILCODE) previously.^10^ The study was registered in ClinicalTrials.gov (Identifier: NCT03370744) and approved by the ethics committee of Xuanwu Hospital of Capital Medical University (Identifier No. [2017]046). The participants were recruited from February 27th to November 15th in 2018, and finally 65 right-handed and Mandarin speaking participants (50 for SCD and 15 for NC) were included. Fig. S1 illustrates the eligibility criteria for the study population.

SCD was defined according to the research criteria for pre-MCI SCD, including 1) self-experienced persistent decline in cognitive capacity compared with a previously normal status and unrelated to an acute event; 2) normal age-, sex-, and education-adjusted performance on standardized cognitive tests and not meet the criteria for MCI or dementia.^1,10^ Individuals with normal performance on the standardized neuropsychological tests and without cognitive problems via the SCD-interview^11^ were included as the NC group.

The exclusion criteria were as follows: 1) a history of stroke; 2) current major psychiatric diagnoses, such as severe depression and anxiety (Hamilton depression scale (HAMD) score > 24 or Hamilton anxiety (HAMA) scale > 29); 3) other diseases that could cause cognitive decline; 4) history of psychosis or congenital mental developmental delay; 5) contraindications for MRI or unable to complete the study protocol.

### Neuropsychological assessments

The neuropsychological tests included three cognitive domains: auditory verbal learning test-Huashan version (AVLT-H), animal fluency test (AFT), 30-item Boston naming test (BNT), and shape trail test (STT) parts A and B.^12-15^ In addition, all subjects implemented tests for global cognition, daily life ability, and neuropsychiatric assessments, including Montreal cognitive assessment-basic (MoCA-B),^16^ functional activities questionnaire (FAQ), HAMA, and HAMD.

### Image data acquisition

For each participant, PET and MRI data were simultaneously acquired with an integrated 3.0 Tesla TOF PET/MR scanner (SIGNA PET/MR, GE Healthcare, Milwaukee, WI, USA) at Xuanwu Hospital of Capital Medical University. Participants were instructed to keep their eyes closed, relax but not fall asleep, and move as little as possible during scanning. Foam pads were used to minimize head movements and earplugs were used to reduce machine noise.

#### PET data acquisition

Participants were scanned with [^18^F] florbetapir (AV45) and FDG-PET in a 3D acquisition mode about four weeks apart (24.4 ± 23.8 days). For FDG-PET, participants were instructed to fast for at least 6□hours. The imaging parameters were listed in the Method-SA.

#### MRI data acquisition

We obtained two fMRI datasets for each participant, namely AV45-fMRI and FDG-fMRI. The detailed scanning parameters for the 3D-T1 and RS-fMRI images were in the Method-SA.

### Data analysis

AV45-PET and FDG-PET images were analyzed by Statistical Parametric Mapping version 12 (SPM 12; https://www.fil.ion.ucl.ac.uk/spm/software/spm12) and in-house software with MATLAB (https://www.mathworks.com). The preprocessing and calculation of the standardized uptake value ratio (SUVR) of the AV45-PET (voxel-wise and global SUVR) and FDG-PET (voxel-wise SUVR) were listed in the Method-SA.

RS-fMRI datasets were preprocessed using the Data Processing Assistant for Brain Imaging (DPABI_V4.1) and Resting-State fMRI Data Analysis Toolkit (RESTplus_1.22).^17,18^ The details were listed in the Method-SA.

Then, three RS-fMRI metrics for local activities were calculated, including ALFF reflecting the amplitude of the fluctuation of each time series of the fMRI signal, ReHo reflecting the local synchronization of the time series of neighboring voxels, and DC reflecting the functional connectivity density of a voxel with all other voxels.^3-5^ It should be noted that, DC is actually an FC index, but often taken as a local metric in graph theory analysis.

Seed-based FC analysis was a post-hoc analysis. As shown in the Results section, 6 voxels in the left dorsal precuneus (dPCu), which showed abnormal activities in 7 of the total 8 neuroimaging metrics in SCD subjects, were taken as a region of interest (named dPCu-ROI hereafter). The details for FC analysis were listed in the Method-SA.

### Statistical analyses

SPSS (version 24.0, IBM) was used for comparisons of demographic and neuropsychological data. DPABI_V4.1 was used for statistical analysis of the local activities and FC between SCD and NC. In addition, the Aβ positivity effect was also tested.^19^ The details were listed in the Method-SA.

## Results

### Behavioral measures

No significant difference was found in the age, sex, or education level between the SCD group and NC group (all *P* > 0.05) (Table 1). The HAMA and HAMD were significantly higher in the SCD group than in the NC group.

**Table 1.**
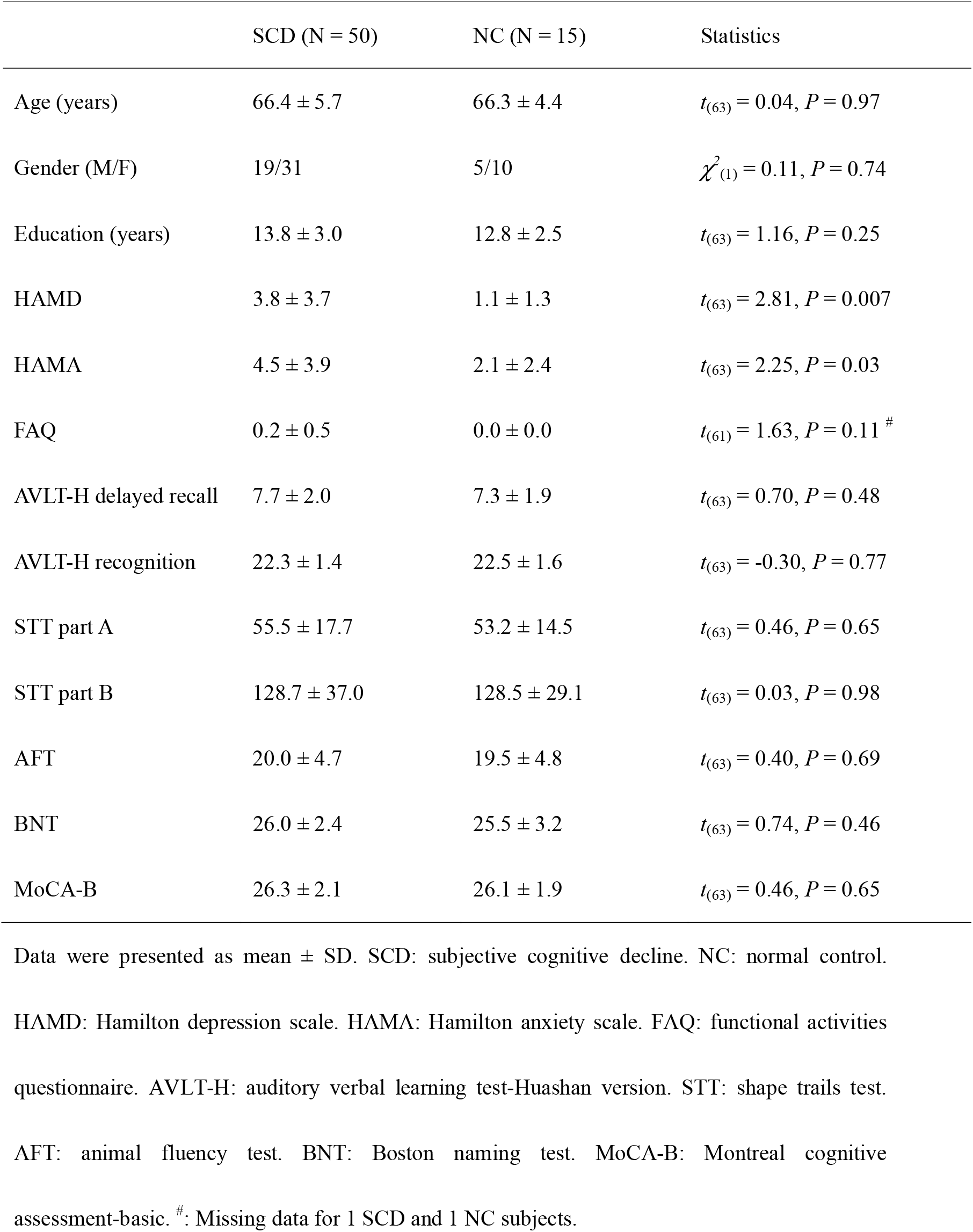
Demographic information and neuropsychological tests.

|  | SCD (N = 50) | NC (N = 15) | Statistics |
| --- | --- | --- | --- |
| Age (years) | 66.4 ± 5.7 | 66.3 ± 4.4 | $t_{(63)} = 0.04, P = 0.97$ |
| Gender (M/F) | 19/31 | 5/10 | $\chi^2_{(1)} = 0.11, P = 0.74$ |
| Education (years) | 13.8 ± 3.0 | 12.8 ± 2.5 | $t_{(63)} = 1.16, P = 0.25$ |
| HAMD | 3.8 ± 3.7 | 1.1 ± 1.3 | $t_{(63)} = 2.81, P = 0.007$ |
| HAMA | 4.5 ± 3.9 | 2.1 ± 2.4 | $t_{(63)} = 2.25, P = 0.03$ |
| FAQ | 0.2 ± 0.5 | 0.0 ± 0.0 | $t_{(61)} = 1.63, P = 0.11$ <sup>#</sup> |
| AVLT-H delayed recall | 7.7 ± 2.0 | 7.3 ± 1.9 | $t_{(63)} = 0.70, P = 0.48$ |
| AVLT-H recognition | 22.3 ± 1.4 | 22.5 ± 1.6 | $t_{(63)} = -0.30, P = 0.77$ |
| STT part A | 55.5 ± 17.7 | 53.2 ± 14.5 | $t_{(63)} = 0.46, P = 0.65$ |
| STT part B | 128.7 ± 37.0 | 128.5 ± 29.1 | $t_{(63)} = 0.03, P = 0.98$ |
| AFT | 20.0 ± 4.7 | 19.5 ± 4.8 | $t_{(63)} = 0.40, P = 0.69$ |
| BNT | 26.0 ± 2.4 | 25.5 ± 3.2 | $t_{(63)} = 0.74, P = 0.46$ |
| MoCA-B | 26.3 ± 2.1 | 26.1 ± 1.9 | $t_{(63)} = 0.46, P = 0.65$ |

Data were presented as mean ± SD. SCD: subjective cognitive decline. NC: normal control.
HAMD: Hamilton depression scale. HAMA: Hamilton anxiety scale. FAQ: functional activities questionnaire. AVLT-H: auditory verbal learning test-Huashan version. STT: shape trails test.
AFT: animal fluency test. BNT: Boston naming test. MoCA-B: Montreal cognitive assessment-basic. <sup>#</sup>: Missing data for 1 SCD and 1 NC subjects.

### Alteration of local activity

The SCD group showed: 1) increased Aβ deposition in the left dPCu, left cuneus, left superior parietal cortex, and right medial superior frontal cortex (Figure 1. *P* < 0.05, FDR corrected); 2) increased glucose metabolism in the left dPCu, left middle occipital cortex, and bilateral paracentral lobule (Figure 1. *P* < 0.05, FDR corrected). The left dPCu was the only region showing overlapped abnormality for Aβ deposition and glucose metabolism.

**Figure 1.**
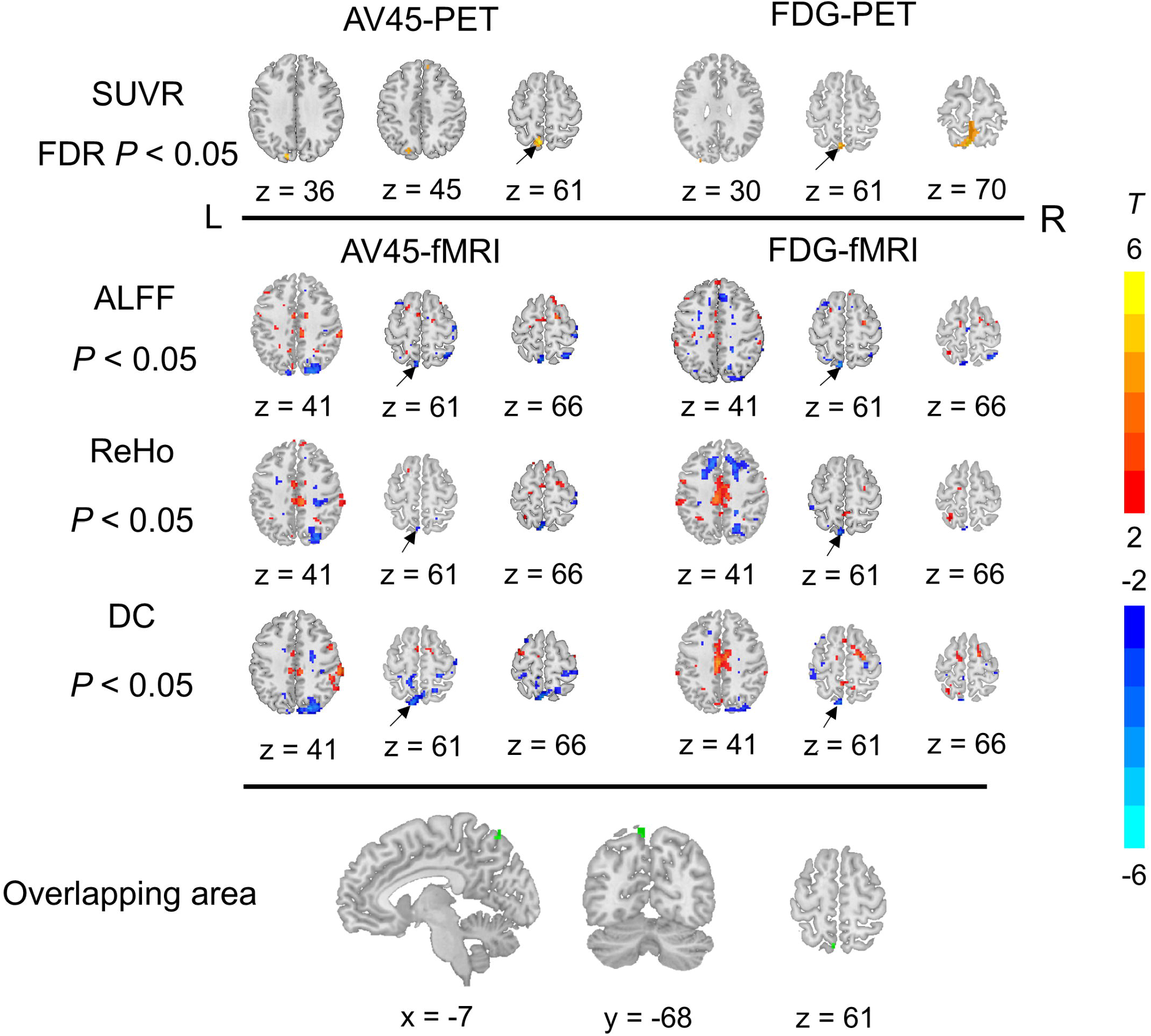
Group differences between subjective cognitive decline (SCD) and normal control (NC) groups with controlling for age, sex, and years of education, on Aβ deposition and glucose metabolism (top row), RS-fMRI metrics (middle row), and the overlapping area of the PET and fMRI metrics (bottom row). Overlapping area at the left dPCu means that 7 of 8 metrics showed group difference, i.e., AV45-SUVR, FDG-SUVR, AV45-ALFF, FDG-ALFF, AV45-ReHo, FDG-ReHo, AV45-DC, and FDG-DC. Red and yellow colors indicate increased metrics in SCD vs. NC, and blue color indicates the opposite. SUVR: standardized uptake value ratio. FDR: false discovery rate. ALFF: amplitude of low frequency fluctuation. ReHo: regional homogeneity. DC: degree centrality. L and R: left and right in the brain. *T*: the *T* value of two-sample *t*-test on the PET and fMRI metrics maps.

The difference of RS-fMRI metrics (ALFF, ReHo, and DC) did not survive FDR correction. With an uncorrected *P* < 0.05, the SCD group showed decreased ALFF, ReHo, and DC in the left dPCu (Figure 1) for both the AV45-fMRI and FDG-fMRI.

### Seed-based functional connectivity

One sample *t*-tests showed that the connectivity patterns of the left dPCu were very similar for AV45-fMRI and FDG-fMRI datasets (*P* < 0.05, FDR corrected, Figure 2). Significantly negative functional connectivity with the left dPCu was mainly in the DMN regions, including the bilateral medial prefrontal cortex, posterior cingulate cortex, and its adjacent ventral PCu, etc. Significantly positive functional connectivity was located in the bilateral inferior frontal cortex, middle frontal cortex, lingual gyrus, etc. No significant difference in RSFC was found between the SCD group and NC group (*P* < 0.05, FDR corrected).

**Figure 2.**
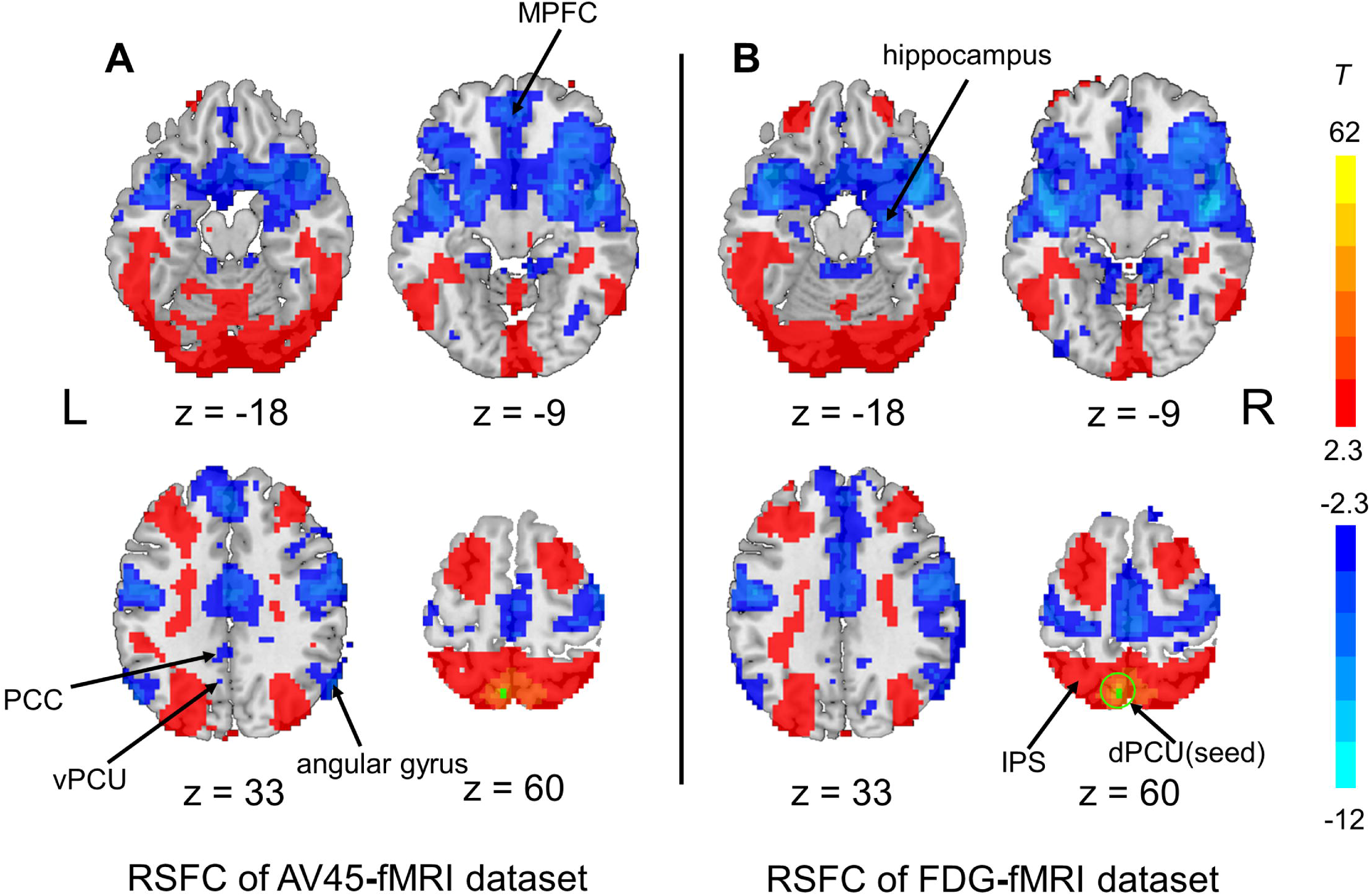
Patterns of seed-based resting-state functional connectivity (RSFC) of the left dorsal precuneus (dPCu) of all the subjects in the AV45-fMRI dataset (A) and FDG-fMRI dataset (B) (P < 0.05, FDR corrected). The yellow-red color indicates positive connections and the blue color indicates negative connections with the left dPCu. MPFC: medial prefrontal cortex. PCC: posterior cingulate cortex. vPCu: ventral PCu. IPS: intraparietal sulcus. L and R: left and right in the brain. *T*: the *T* value of one-sample *t*-test on the RSFC maps.

### Correlation between imaging metrics and behavioral measures in the SCD group

We observed significant correlation between any pair of two fMRI metrics (ALFF, ReHo, and DC; *P* < 0.001) within the dPCu-ROI. No significant correlation was found between other pairs of imaging metrics (Table S2).

As for the correlation between behavioral scales and imaging metrics, although a few correlation showed a *P* value between 0.05 – 0.01, none survided multiple comarision correction (Table S3).

### Aβ deposition positivity and neuroimaging metrics in the dPCu-ROI

The demographic and neuropsychological data of NC^-^, SCD^-^ and SCD^+^ groups can be found in Table S4. The HAMD score was significantly higher in the SCD^+^ group and the SCD^-^ group than in the NC^-^ group.

SCD^+^ group showed significantly higher global AV45-SUVR than both NC^-^ and SCD^-^ groups, but no significant difference was found between NC^-^ and SCD^-^ (*P* = 0.43) (Figure 3A). Both SCD^-^ and SCD^+^ groups had significantly higher AV45-SUVR in left dPCu-ROI than NC-group, while no significant difference was found between SCD^-^ and SCD^+^ (*P* = 0.08) (Figure 3B).With regard to the FDG-SUVR and all RS-fMRI metrics, both SCD^+^ and SCD^-^ showed hypermetabolism and decreased fMRI activity in the dPCu-ROI, except for the comparison of FDG-ReHo between SCD^+^ group and NC^-^ group (*P* = 0.11) (Figure 3).

**Figure 3.**
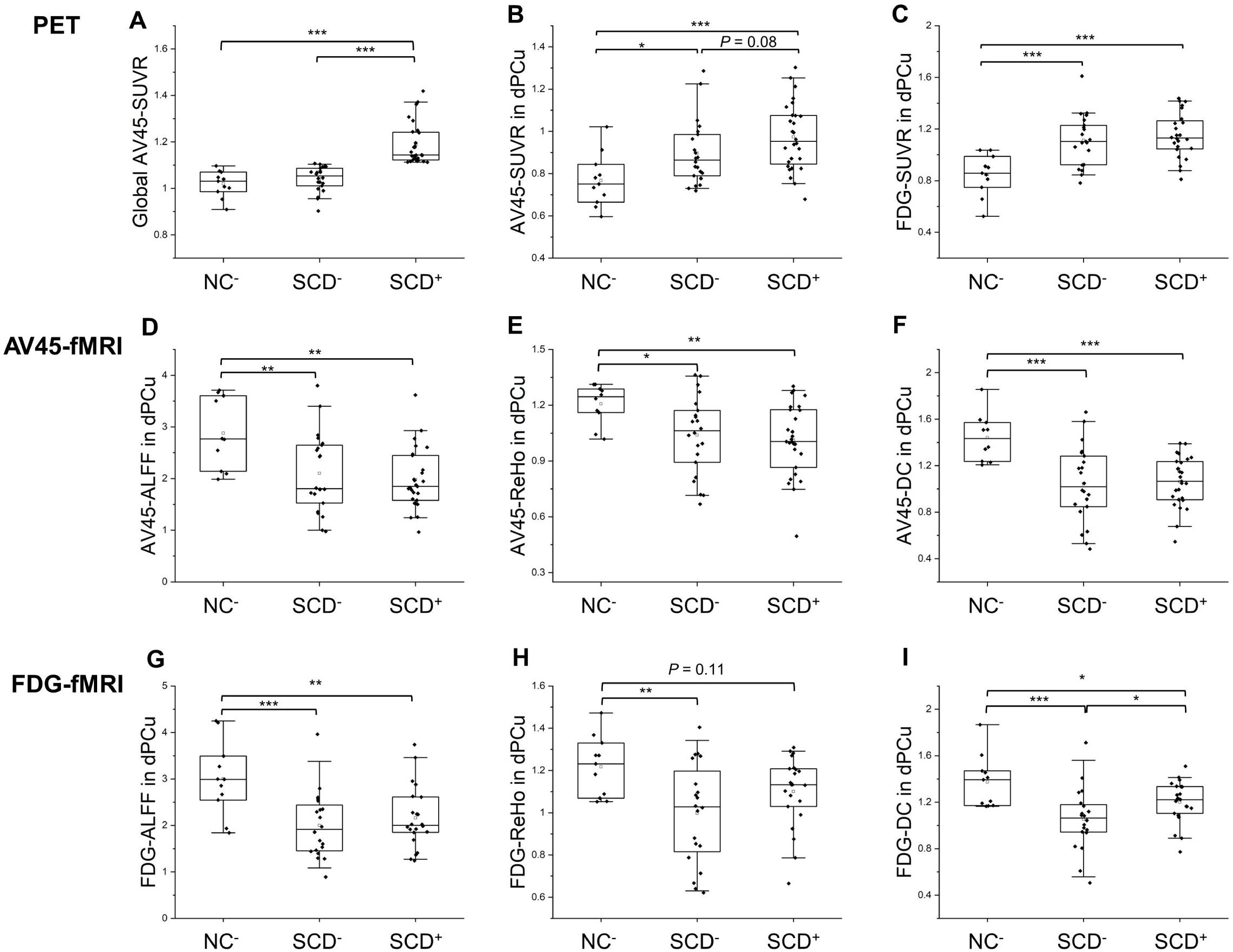
Comparisons of global AV45-SUVR (A) and the 8 metrics in the left dPCu-ROI among three groups, i.e. AV45-SUVR (B), FDG-SUVR (C), AV45-ALFF (D), AV45-ReHo (E), AV45-DC (F), FDG-ALFF (G), FDG-ReHo (H), and FDG-DC (I). NC^-^: Aβ negative NC. SCD^-^: Aβ negative SCD. SCD^+^: Aβ positive SCD. * : *P* < 0.05, ** : *P* < 0.01, *** : *P* < 0.001. All box and whisker plots: box range, 25–75%; whisker range, 5–95%.

### Independent verification of data analyses

In order to controlling for the potential influence from data anslysis upon our results, we replicated all the analyses by two independent research teams based on the same software and pipeline. The results were substaitially the same (Fig. S2-4 and Table S5-6).

## Discussion

### Convergent abnormal local activity in AD/MCI/SCD

In the PCC and its adjacent PCu (sometimes named PCC/PCu) of MCI and AD, meta-analyses have reported increased Aβ deposition and decreased glucose metabolism, decreased RS-fMRI ALFF, and ReHo.^6,8,9^ However, the current study found convergent abnormal activities in 7/8 functional metrics in the dPCu. Furthermore, the seed-based RS-fMRI FC showed that the dPCu was negatively connected with the DMN (including PCC and its adjacent ventral PCu). Previous RS-fMRI studies have segmented the precuneus into 6 or 8 sub-regions,^20,21^ suggesting precuneus may be functionally heterogeneous. We searched in the Neurosynth (http://neurosynth.org) (coordinates: −6, −68, 62; distance: 4 mm) and found that the dPCu was reported to be involved in more cognitive than memory tasks, e.g., unfocused attention,^22^ in healthy people. Although many task fMRI studies have been carried out on SCD (Table S7), none has reported abnormal activation in the dPCu. Future studies should investigate the specific cognitive processing of the dPCu, and hence develop more specific cognitive tests for SCD.

### Increased Aβ deposition vs. increased glucose metabolism in SCD

Meta-analysis has reported that the increased Aβ deposition was associated with decreased glucose metabolism in the same area of PCC in MCI/AD,^6^ indicating a decreased neuronal activity. However, we found an association of increased Aβ deposition with increased glucose metabolism in the dPCu, indicating an increased activity. No previous PET imaging studies of SCD, MCI and AD reported one positive relationship between cerebral Aβ deposition and glucose metabolism as we found in this study. A few human PET and task fMRI studies^23-25^ indirectly support our findingsStudies of animal model of early AD have frequently reported Aβ-dependent increased neuronal activity.^26,27^ Our results support the view that Aβ-dependent hyperactivity may occur long before the presence of AD symptoms.^26^

### Decreased value of RS-fMRI local metrics vs. increased glucose metabolism in SCD

As aforementioned, for MCI and AD, both the decreased glucose metabolism and decreased value of RS-fMRI local metrics (ALFF and ReHo) in the PCC were interpreted as decreased neuronal activity. However, we found seemingly discrepant results, i.e., increased glucose metabolism and decreased value of the local RS-fMRI metrics in the dPCu. The following notations might be helpful.

1. Mathematically, the glucose metabolism of FDG-PET reflects the integrated or mean value over a period of time, while the RS-fMRI ALFF reflects the amplitude of fluctuation or standard deviation within a specific frequency band. The simulation result showed that the mean value has no linear correlation with the standard deviation (Fig. S5). It has commonly been suggested that the fMRI signal is more coupled with the local field potential (LFP) than the action potential.^28^ Mathematically, both the RS-fMRI signal and the LFP are analyzed in a frequency-dependent way, however, their frequency-dependent correlation of the amplitude, a popular metric for both RS-fMRI and LFP, is far from clear.
2. A few PET-fMRI studies have shown that, although both the glucose metabolism^29^ and RS-fMRI local metrics show similar higher activity in the default mode network (DMN) regions (e.g., PCC) than other brain regions, voxel-wise across-subject correlation did not show robust significant linear correlation (Table S8).^30-32^
3. Brain disorders may show opposite association of glucose metabolism with RS-fMRI local metrics, e.g., increased glucose metabolism^33^ but decreased ALFF^34^ in the putamen of patients with Parkinson’s disease (Table S9). The current study also showed opposite association in the dPCu, however, as aforementioned, AD/MCI showed positive association, i.e., decreased glucose metabolism and decreased value of RS-fMRI metrics.^6,8,9^

Considering the non-invasiveness, low-cost, and easy-accessing advantages of RS-fMRI technique, it may be a promising technique for precise localizing the AD-related abnormal activity, and hence, could be used to guide precise brain stimulation, e.g., transcranial magnetic stimulation.

### SCD showed abnormal local activities regardless of Aβ positivity

Binary classification of Aβ deposition into positive and negative is a popular way for clinical diagnosis for either healthy aging or participants with SCD.^35^ The current study thus classified all participants into SCD^+^, SCD^-^, NC^+^, and NC^-^ with the criteria of the cortical summary AV45-SUVR value ≥ 1.11.^19^ NC^+^ group was not included into the further analysis due to the limited sample size (n = 4). Both the SCD^-^ and SCD^+^ groups showed increased Aβ deposition and glucose metabolism and decreased value of RS-fMRI metrics in the dPCu as compared with NC^-^ group. In other words, like the SCD^+^ group, the SCD^-^ group also showed evidences of elevated Aβ pathology and hypermetabolism, as well as abnormal fMRI activities. Although the Aβ +/- categorization is often used to define preclinical AD, it does not imply that gobally Aβ negative subjects have no pathology in the brain.^35,36^ Consistent with our findings, previous literature also found that local regions (including precuneus and posterior cingulate etc.) may be more sensitive to detect early Aβ deposition than global cortical summary regions.^37-39^ Furthermore, we also compared the mean values of the eight metrics in the PCC (Fig. S6), the SCD^+^ group showed significantly higher AV45-SUVR than the SCD^-^ and NC^-^ group. However, we did not found significant difference in the FDG-SUVR and fMRI metrics of PCC between the NC-group and the two SCD groups. Together, it is likely that the increased Aβ deposition may occur concurrently with hypermetabolism and abnormal fMRI activities in dPCu (early Aβ-affected region) in the early stage of AD.

### Limitations

This study has some limitations. First, this is a cross-sectional data, thus further longitudinal studies would be helpful to investigate the changes of Aβ deposition, neuronal dysfunction, and cognitive function as the disease progresses. Second, we did not have tau PET imaging, thus we can not evaluate tau pathology in this study. Third, although multimodal metrics and two fMRI datasets were used to increase the reliability of our results, data from multiple research centers are needed for validation of the findings in this study. Finally, this study did not include cognitive tests related to unfocused attention which might be more specific to elaborate the functional roles of the dPCu.

## Conclusions

In summary, we found increased Aβ deposition, increased glucose metabolism, and decreased value of RS-fMRI metrics in the same area of the left dPCu in SCD. The negative functional connectivity between the left dPCu and DMN suggests that the dPCu is not a region in the DMN. The left dPCu may be among the earliest impaired regions, and the DMN is probably impaired in a later stage, e.g., MCI/AD. Precisely targeting on the left dPCu, future studies should develop delicate cognitive tasks to probe the cognitive impairments by task fMRI and/or virtual lesion of transcranial magnetic stimulation (TMS). Further, repetitive TMS could be used as an early intervention on the dPCu.

## Supporting information

checklist

## Data Availability

The dataset generated and analyzed in the current study is available from the corresponding author on reasonable request.

## Funding

This work was supported by the National Natural Science Foundation of China (61633018, 82020108013 and 81661148045).

## Data Availability Statement

The dataset generated and analyzed in the current study is available fromthe corresponding author on reasonable request.

## Conflict of Interest

The authors declare that the research was conducted in the absence of any commercial or financial relationships that could be construed as a potential conflict of interest.

## Notes

### Competing Interest Statement

The authors have declared no competing interest.

### Clinical Trial

NCT03370744

### Author Declarations

The ethics committee of Xuanwu Hospital of Capital Medical University.

## References

1. Jessen F, Amariglio RE, van Boxtel M, et al. A conceptual framework for research on subjective cognitive decline in preclinical Alzheimer’s disease. Alzheimers Dement 2014;10:844–852.

2. Molinuevo JL, Rabin LA, Amariglio R, et al. Implementation of subjective cognitive decline criteria in research studies. Alzheimers Dement 2017;13:296–311.

3. Zang YF, He Y, Zhu CZ, et al. Altered baseline brain activity in children with ADHD revealed by resting-state functional MRI. Brain Dev 2007;29:83–91.

4. Zang Y, Jiang T, Lu Y, He Y, Tian L. Regional homogeneity approach to fMRI data analysis. Neuroimage 2004;22:394–400.

5. Zuo XN, Ehmke R, Mennes M, et al. Network centrality in the human functional connectome. Cereb Cortex 2012;22:1862–1875.

6. He W, Liu D, Radua J, Li G, Han B, Sun Z. Meta-analytic comparison between PIB-PET and FDG-PET results in Alzheimer’s disease and MCI. Cell Biochem Biophys 2015;71:17–26.

7. Reiman EM, Caselli RJ, Yun LS, et al. Preclinical evidence of Alzheimer’s disease in persons homozygous for the epsilon 4 allele for apolipoprotein E. N Engl J Med 1996;334:752–758.

8. Pan P, Zhu L, Yu T, et al. Aberrant spontaneous low-frequency brain activity in amnestic mild cognitive impairment: A meta-analysis of resting-state fMRI studies. Ageing Res Rev 2017;35:12–21.

9. Zheng D, Xia W, Yi ZQ, et al. Alterations of brain local functional connectivity in amnestic mild cognitive impairment. Transl Neurodegener 2018;7:26.

10. Li X, Wang X, Su L, Hu X, Han Y. Sino longitudinal study on cognitive decline (SILCODE): protocol for a Chinese longitudinal observational study to develop risk prediction models of conversion to mild cognitive impairment in individuals with subjective cognitive decline. BMJ Open 2019;9:e028188.

11. Jessen F, Spottke A, Boecker H, et al. Design and first baseline data of the DZNE multicenter observational study on predementia Alzheimer’s disease (DELCODE). Alzheimers Res Ther 2018;10:15.

12. Zhao Q, Lv Y, Zhou Y, Hong Z, Guo Q. Short-term delayed recall of auditory verbal learning test is equivalent to long-term delayed recall for identifying amnestic mild cognitive impairment. PLoS One 2012;7:e51157.

13. Guo Q, Jin L, Hong Z, Lv C. A specific phenomenon of animal fluency test in Chinese elderly. Chinese Mental Health Journal 2007;21:622–625.

14. Guo Q, Hong Z, Shi W, Sun Y, Lv C. Boston naming test in Chinese elderly, patient with mild cognitive impairment and Alzheimer’s dementia. Chinese Mental Health Journal 2006;20:81–84.

15. Zhao Q, Guo Q, Li F, Zhou Y, Wang B, Hong Z. The Shape Trail Test: application of a new variant of the Trail making test. PLoS One 2013;8:e57333.

16. Chen KL, Xu Y, Chu AQ, et al. Validation of the Chinese Version of Montreal Cognitive Assessment Basic for Screening Mild Cognitive Impairment. J Am Geriatr Soc 2016;64:e285–e290.

17. Yan CG, Wang XD, Zuo XN, Zang YF. DPABI: Data Processing & Analysis for (Resting-State) Brain Imaging. Neuroinformatics 2016;14:339–351.

18. Jia XZ, Wang J, Sun HY, et al. RESTplus:an improved toolkit for resting-state functional magnetic resonance imaging data processing. Sci bull 2019;64:953–954.

19. Landau SM, Breault C, Joshi AD, et al. Amyloid-β imaging with Pittsburgh compound B and florbetapir: comparing radiotracers and quantification methods. J Nucl Med 2013;54:70–77.

20. Luo Z, Zeng LL, Qin J, Hou C, Shen H, Hu D. Functional Parcellation of Human Brain Precuneus Using Density-Based Clustering. Cereb Cortex 2020;30:269–282.

21. Zhang S, Li CS. Functional connectivity mapping of the human precuneus by resting state fMRI. Neuroimage 2012;59:3548–62.

22. Dickenson J, Berkman ET, Arch J, Lieberman MD. Neural correlates of focused attention during a brief mindfulness induction. Soc Cogn Affect Neurosci 2013;8:40–47.

23. Cohen AD, Price JC, Weissfeld LA, et al. Basal cerebral metabolism may modulate the cognitive effects of Abeta in mild cognitive impairment: an example of brain reserve. J Neurosci 2009;29:14770–14778.

24. Oh H, Madison C, Baker S, Rabinovici G, Jagust W. Dynamic relationships between age, amyloid-β deposition, and glucose metabolism link to the regional vulnerability to Alzheimer’s disease. Brain 2016;139:2275–2289.

25. Sperling RA, Dickerson BC, Pihlajamaki M, et al. Functional alterations in memory networks in early Alzheimer’s disease. Neuromolecular Med 2010;12:27–43.

26. Zott B, Busche MA, Sperling RA, Konnerth A. What happens with the circuit in Alzheimer’s disease in mice and humans? Annu Rev Neurosci 2018;41:277–297.

27. Zott B, Simon MM, Hong W. A vicious cycle of β amyloid-dependent neuronal hyperactivation. Science 2019;365:559–565.

28. Logothetis NK, Pauls J, Augath M, Trinath T, Oeltermann A. Neurophysiological investigation of the basis of the fMRI signal. Nature 2001;412:150–157.

29. Raichle ME, MacLeod AM, Snyder AZ, Powers WJ, Gusnard DA, Shulman GL. A default mode of brain function. Proc Natl Acad Sci U S A 2001;98(2):676–682.

30. Aiello M, Salvatore E, Cachia A, et al. Relationship between simultaneously acquired resting-state regional cerebral glucose metabolism and functional MRI: A PET/MR hybrid scanner study. Neuroimage 2015;113:111–121.

31. Bernier M, Croteau E, Castellano CA, Cunnane SC, Whittingstall K. Spatial distribution of resting-state BOLD regional homogeneity as a predictor of brain glucose uptake: A study in healthy aging. Neuroimage 2017;150:14–22.

32. Jiao F, Gao Z, Shi K, et al. Frequency-dependent relationship between resting-state fMRI and glucose metabolism in the elderly. Front Neurol 2019;10:566.

33. Tomše P, Jensterle L, Grmek M, et al. Abnormal metabolic brain network associated with Parkinson’s disease: replication on a new European sample. Neuroradiology 2017;59:507–515.

34. Wang J, Zhang JR, Zang YF, Wu T. Consistent decreased activity in the putamen in Parkinson’s disease: a meta-analysis and an independent validation of resting-state fMRI. Gigascience 2018;7:giy071.

35. Jack CR Jr, Bennett DA, Blennow K, et al. A/T/N: An unbiased descriptive classification scheme for Alzheimer disease biomarkers. Neurology 2016;87:539–47.

36. Jack CR Jr, Bennett DA, Blennow K, et al. NIA-AA Research Framework: Toward a biological definition of Alzheimer’s disease. Alzheimers Dement 2018;14:535–562.

37. Palmqvist S, Schöll M, Strandberg O, et al. Earliest accumulation of β-amyloid occurs within the default-mode network and concurrently affects brain connectivity. Nat Commun 2017;8:1214.

38. Farrell ME, Chen X, Rundle MM, Chan MY, Wig GS, Park DC. Regional amyloid accumulation and cognitive decline in initially amyloid-negative adults. Neurology 2018;91:e1809–e1821.

39. Guo T, Landau SM, Jagust WJ; Alzheimer’s Disease Neuroimaging Initiative. Detecting earlier stages of amyloid deposition using PET in cognitively normal elderly adults. Neurology 2020;94:e1512–e1524.

