## Supplementary Appendix for "Convergent abnormalities of β-amyloid deposition, glucose metabolism, and fMRI activity in the dorsal precuneus in subjective cognitive decline"

**List of investigators**

Xuanwu Hospital of Capital Medical University, Department of Neurology (Xuan-Yu Li, Wen-Ying Du, Ying Han, Wei-Na Zhao, Zi-Qi Wang, Ying Chen, Yu-Xia Li, Bin Mu, Xing Zhao, Jun Wang, Ming-Yan Zhao, Liu Yang, Guan-Qun Chen, Qin Yang, Jia-Chen Li, Xiao-Qi Wang, Li Lin, Yu Sun, Xiao-Ni Wang, Can Sheng, Tao-Ran Li, Yi Liu); Hangzhou Normal University, The Affiliated Hospital of Hangzhou Normal University, Center for Cognition and Brain Disorders (Li-Xia Yuan, Yu-Feng Zang); Shanghai University, Department of Shanghai Institute for Advanced Communication and Data Science (Chang-Chang Ding, Jie-Hui Jiang); Shenzhen Bay Laboratory, Institute of Biomedical Engineering (Teng-Fei Guo); University of Cologne, Medical Faculty, Department of Psychiatry (Frank Jessen); Capital Medical University, Beijing Friendship Hospital, Department of Neurology (Xuan-Yu Li); Zhejiang Key Laboratory for Research in Assessment of Cognitive Impairments (Li-XiaYuan, Yu-Feng Zang); Hangzhou Normal University, Center for Cognition and Brain Disorders and the Affiliated Hospital (Li-XiaYuan, Yu-Feng Zang); Beijing Institute for Brain Disorders, Center of Alzheimer’s Disease (Ying Han); National Clinical Research Center for Geriatric Disorders (Ying Han); Hainan University, School of Biomedical Engineering (Fei-Fan Zhou, Ying Han).

**Methods**

1. **Image data acquisition**
   1.1 **PET data acquisition**

Participants were scanned with [^18^F] florbetapir (AV45) and [^18^F]-fluorodeoxyglucose (FDG)-PET in a 3D acquisition mode about four weeks apart (24.4 ± 23.8 days). For FDG-PET, participants were instructed to fast for at least 6 hours. A static scan of 10 minutes was performed 40 to 60 minutes after an intravenous injection of 3.7 MBq/kg of [^18^F] FDG. For AV45-PET, a static scan of 10 minutes was acquired 40 to 60 minutes after an intravenous injection of 7-10 mCi [^18^F] florbetapir. All the PET data were reconstructed using a time-of-flight ordered subset expectation maximization (TOF-OSEM) algorithm with the following parameters: 8 iterations, 32 subsets, matrix size = 192 × 192, field of view (FOV) = 350 × 350 mm^2^, Gaussian filter (full width at half maximum (FWHM) 3 mm), and correction for the attenuation, scatter, random counts, and dead-time.

1.2 **MRI data acquisition**

We obtained two fMRI datasets for each participant, namely AV45-fMRI and FDG-fMRI. RS-fMRI images were acquired with a single-shot gradient-echo-planar imaging sequence, using the following parameters: repetition time (TR) = 2000 ms, echo time (TE) = 30 ms, flip angle = 90°, slice thickness =3.0 mm, gap = 1.0 mm, FOV = 224 × 224 mm^2^, data matrix = 64 × 64, slice number = 28, voxel size = 3.5 × 3.5 × 4 mm^3^, interleaved slice order, 240 volumes (8 min) in total. The detailed scanning parameters for the 3D-T1 spoiled gradient recalled sequence were: TR = 6.9 ms, TE = 2.98 ms, inversion time = 450 ms, flip angle = 12°, matrix size = 256 × 256 × 192, voxel size = 1 × 1 × 1 mm^3^.

2. Data analysis

**2.1 PET data processing**

Four subjects were excluded due to quality issue of AV45-PET images (Fig. S7). AV45-PET and FDG-PET images were preprocessed by Statistical Parametric Mapping version 12 (SPM 12; <https://www.fil.ion.ucl.ac.uk/spm/software/spm12>) and in-house software with MATLAB (https://www.mathworks.com). The 3D-T1 images were firstly registrated to PET images and segmented.^1^ Then, the PET images were spatially normalized to Montreal Neurological Institute (MNI) standard space with the forward deformation field estimated during the unified segmentation, resampled to an isotropic voxel size of 3 × 3 × 3 mm^3^, and smoothed with a Gaussian kernel of 10 mm FWHM. Finally, the voxel-wise standardized uptake value ratio (SUVR) were obtained by choosing the whole cerebellum (for AV45-PET) and pons and vermis (for FDG-PET) as the reference region.^2^ The global AV45-SUVR was computed by averaging the SUVR in a set of regions including frontal, temporal, parietal and cingulate cortices.^3^

**2.2. RS-fMRI data processing**

RS-fMRI datasets were preprocessed using the Data Processing Assistant for Brain Imaging (DPABI_V4.1) and Resting-State fMRI Data Analysis Toolkit (RESTplus_1.22).^4,5^ The first 10 volumes were discarded to have the participants get used to the scanning noise. The remaining volumes were corrected for slice time differences and then realigned to the first volume for motion correction. Next, the 3D-T1 images were coregistered to the averaged functional image and segmented. The motion-corrected functional volumes were spatially normalized to MNI standard space with the forward deformation field from unified segmentation and resampled to 3 × 3 × 3 mm^3^. The resulting images were spatially smoothed with a Gaussian kernel of 6 mm FWHM. It should be noted that the spatial smoothing was performed before the calculation of ALFF and seed-based RSFC, while after the calculation of ReHo and DC. Then, the linear trend of time courses was removed. Nuisance signals (including Friston-24 head motion parameters, white matter signal, and cerebrospinal fluid signal) were extracted and regressed out from the data. Finally, a band-pass filtering (0.01 – 0.08 Hz) was performed for before ReHo and DC calculation, but not for ALFF calculation.

ALFF analysis: Before ALFF analysis, spatial smoothing (Gaussian kernel with 6 mm FWHM) was performed. The time course of each voxel was transformed into the frequency domain with the fast Fourier transform. The square root of the power spectrum was calculated and averaged across 0.01 - 0.08 Hz.^6^ This averaged square root was the ALFF value for a given voxel. For standardization, each voxel’s ALFF value was divided by the global mean ALFF, and hence an mALFF map was obtained.

ReHo analysis: Kendall’s coefficient of concordance (KCC) was computed on the ranked time course of a given voxel and its 26 nearest neighbors.^7^ The obtained KCC was the ReHo value. Then, mReHo maps were obtained as mALFF maps.

DC analysis: DC represents the functional connection strength between a given voxel and all the voxels in the brain.^8^ First, we computed the Pearson correlation between the time course of a given voxel and all the other voxels in the whole brain. Then, the Pearson correlation coefficients larger than 0.2 were summed and the then assigned to that given voxel as DC value. Standardized mDC maps were created as mALFF.

RSFC analysis: The RSFC was a post-hoc analysis. First, the global mean signal was extracted and regressed out from the data. As shown in the Results section (Figure 1), an area in the left dorsal PCu (6 voxels), which showed abnormal activity in 7 of the total 8 neuroimaging metrics in SCD subjects, was taken as a region of interest (ROI, named dorsal PCu ROI or dPCu-ROI hereafter). Individual RSFC maps of the dPCu-ROI were generated by calculating Pearson’s correlation coefficients between the mean time course of the dPCu-ROI and the time course of each voxel in the whole brain. Subject-level RSFC maps were then converted to z-value maps (zRSFC) with Fisher’s transformation to improve the normality.

3. Statistical analysis

SPSS (version 24.0, IBM) and DPABI_V4.1 were used for statistical analysis.

3.1 **Demographic and neuropsychological data comparison**

Group comparisons on demographic and neuropsychological data were performed with two-sample *t*-test and the Chi-square analyses.

3.2 **Local activity alteration**

Two-sample *t*-tests were conducted between SCD group and NC group on the eight local neuroimaging metrics (AV45-SUVR, FDG-SUVR, AV45-ALFF, AV45-ReHo, AV45-DC, FDG-ALFF, FDG-ReHo, and FDG-DC) with age, gender, and years of education as covariates. False discovery rate (FDR) (*P* < 0.05) was used for multiple comparison correction. The subject numbers in the statistical analyses for each functional neuroimaging modality were summarized in Table S10.

3.3 **Functional connectivity alteration**

One-sample *t-*test was performed on the zRSFC maps of AV45-fMRI and FDG-fMRI datasets for all subjects, respectively. Then, regions showing significant correlation (*P* < 0.05, FDR corrected) entered into two-sample *t*-tests between SCD group and NC group with the age, gender, and years of education as covariates. FDR (*P* < 0.05) was used for multiple comparison correction.

**3.4 Correlation between neuroimaging metrics and behavioral measures**

Partial correlation analysis was performed to investigate the relationship between the eight local neuroimaging metrics (the mean values in the dPCu-ROI of AV45-SUVR, FDG-SUVR, AV45-ALFF, AV45-ReHo, AV45-DC, FDG-ALFF, FDG-ReHo, and FDG-DC) and neuropsychological measures (AVLT-H delayed recall, AVLT-H recognition, STT A and B, AFT, BNT, MoCA-B, HAMD, and HAMA) in the SCD group with age, gender, and education level as covariates.

3.5 **Sub-groups of positivity and negativity of Aβ deposition**

Considering the fact that the Aβ positivity is widely used in clinical practice and research, we thus re-grouped all participants by Aβ positivity. A participant was defined as Aβ positive if the global AV45-SUVR value ≥ 1.11,^3^ we thus got Aβ negative NC (NC^-^, n = 11), Aβ positive NC (NC^+^, n = 4), Aβ negative SCD (SCD^-^, n = 22), and Aβ positive SCD (SCD^+^, n = 28). the NC^+^ group was excluded because of the small sample size (n = 4). Group comparisons on demographic, neuropsychological data, global AV45-SUVR, and mean values of the eight neuroimaging metrics in the dPCu-ROI were performed by one-way ANOVA with least significant difference (LSD) post-hoc test. Furthermore, we also compared the mean values of the eight metrics in the PCC ROI (available for download on the LONI website),^2^ one of the most important abnormal regions that are indicative of pathological change in MCI and AD.

**Supplementary Tables**

**Table S1. Summary of neuroimaging studies on SCD.**

| **Modality** | **References** | **Sample (mean age ± SD)** | **Main findings related to our results** |
| --- | --- | --- | --- |
| Amyloid-PET^a^ |  | | |
|  | Li et al. (2018) | NC, n = 40 (75.10 ± 5.39)  SMC, n = 44 (73.78 ± 5.81) | The voxel-wise comparison of Aβ deposition showed no significant difference between the SMC and NC groups. |
|  | Risacher et al. (2015) | NC ε4-, n = 132 (73.7 ± 6.1)  NC ε4+, n = 53 (71.8 ± 6.4)  SMC ε4-, n = 71 (72.5 ± 5.7)  SMC ε4+, n =33 (70.3 ± 5.2)  EMCI ε4-, n =174 (71.6 ± 7.3)  EMCI ε4+, n = 131 (70.0 ± 7.5) | A main effect of diagnosis was also observed (EMCI, SMC, NC), with significant Aβ deposition seen in primarily the frontal, temporal, and medial parietal cortices. |
|  | Yasuno et al. (2015) | nSCI, n = 30 (72.2 ± 4.8)  SCI, n = 23 (69.6 ± 8.0) | There were no significant differences in PIB-BPND values between the SCI (n = 13) and nSCI (n = 15) in the whole-brain voxel-based analysis. There were 3 Aβ-positive (BPND > 0.25) subjects among the 15 nSCI subjects and 1 among the 13 SCI subjects. |
|  | Snitz et al. (2015) | NC, n = 84 (68.1 ± 4.0)  SCI, n = 14 (73.6 ± 5.8) | SCD participants had higher mean SUVR in frontal cortex, lateral temporal cortex and parietal cortex compared to NC. There were no significant group differences in precuneus, anterior cingulate or anterior ventral striatum. |
| FDG-PET^b^ |  | | |
|  | Matías-Guiu et al. (2017) | NC, n = 20 (65.0 ± 10.6)  SMC, n = 9 (72.4 ± 10.6)  GDS 3, n = 39 (72.9 ± 8.9)  GDS 4, n = 32 (75.5 ± 6.9) | The SMC group did not display any regions with increased or decreased metabolism with respect to those regions in healthy controls. |
|  | Risacher et al. (2015) | NC ε4-, n = 132 (73.7 ± 6.1)  NC ε4+, n = 53 (71.8 ± 6.4)  SMC ε4-, n = 71 (72.5 ± 5.7)  SMC ε4+, n = 33 (70.3 ± 5.2)  EMCI ε4-, n = 174 (71.6 ± 7.3)  EMCI ε4+, n = 131 (70.0 ± 7.5) | A trend toward higher cortical glucose metabolism in SMC ApoE ε4+, although this did not reach significance after Bonferroni-correction. |
|  | Scheef et al. (2012) | NC, n = 56 (66.4 ± 7.2)  SMI, n = 31 (67.6 ± 6.2) | The SMI group showed hypometabolism in the right precuneus, the left parietal cortex (uncorrected statistical threshold), and the left precuneus (ROI analysis) and hypermetabolism in the right medial temporal lobe. |
|  | Rimajova et al. (2008) | SMC, n = 23 (65.05 ± 9.27)  non-SMC, n = 7 (66.74 ± 11.31) | When SMCs were compared to the NeuroStat database, the anterior (unilaterally) and posterior (bilaterally) cingulate regions were shown to manifest a significant reduction in glucose metabolism. No significant changes were observed when non-SMCs were compared to the database, or when non-SMCs were compared to SMC participants. |
|  | Mosconi et al. (2008) | SMC- ε4-, n = 7 (63 ± 5) SMC- ε4+, n = 8 (60 ± 9) SMC+ ε4-, n = 7 (54 ± 9) SMC+ ε4+, n = 6 (59 ± 7) | SMC+ subjects showed reduced parietotemporal and parahippocampal gyrus CMRglc. |
|  | Song et al. (2016) | NC, n = 42 (68.02 ± 5.44) SMI, n = 68 (69.94 ± 6.44) MCI, n = 47 (69.55 ± 6.65) | SMI group showed a significant reduction in glucose metabolism in the periventricular regions. |
|  | Brugnolo et al. (2014) | NC, n = 30 (71.87 ± 7.08) SMC, n = 32 (70.47 ± 7.68) aMCI, n = 22 (74.27 ± 6.66) | No significant difference was found in the controls minus SMC comparisons. |
| Amyloid & FDG-PET^c^ | | | |
|  | Kuhn et al. (2019) | NC, n = 28 (72.25 ± 6.33)  SCD-community, n = 23 (71.70 ± 6.60)  SCD-clinic, n = 27 (68.30 ± 7.99) | In the SCD-community group, self-reported global_R_ SCD negatively correlated with glucose metabolism and grey matter volume in the left insula, right superior frontal, and anterior cingulate cortex. In the SCD-clinic group, the self-reported global_R_ SCD negatively correlated with glucose metabolism in the bilateral insula, left medial prefrontal cortex, bilateral superior and middle temporal cortex, and right fusiform gyrus. |
|  | Amariglio et al. (2015) | Stage 0 (Aβ-/ND-)  Stage 1 (Aβ+/ND-) Stage 2 (Aβ+/ND+) SNAP (Aβ-/ND+) | Both Aβ and ND were independently associated with greater SCC controlling for objective memory performance. Individuals who were biomarker-positive on both Aβ and ND had the highest SCC compared to individuals who were biomarker-positive on either Aβ or ND in isolation. |
|  | Risacher et al. (2015) | NC ε4-, n = 132 (73.7 ± 6.1)  NC ε4+, n = 53 (71.8 ± 6.4)  SMC ε4-, n = 71 (72.5 ± 5.7)  SMC ε4+, n = 33 (70.3 ± 5.2)  EMCI ε4-, n = 174 (71.6 ± 7.3)  EMCI ε4+, n = 131 (70.0 ± 7.5) | A main effect of diagnosis was also observed (EMCI, SMC, NC), with significant Aβ deposition seen in primarily the frontal, temporal, and medial parietal cortices. A trend toward higher cortical glucose metabolism in SMC APOE ε4+, although this did not reach significance after Bonferroni-correction. |
|  | Wirth et al. (2017) | NC, n = 41 (66.1 ± 7.7) APOE4+, n = 17 (63.9 ± 8.6) SCD, n = 16 (68.9 ± 7.3) MCI, n = 30 (73.4 ± 7.2) AD, n = 22 (68.7 ± 9.4) | In SCD patients, Aβ load predominated over local levels of GMV or metabolism within local clusters in prefrontal and posterior cingulate regions, respectively. There was no region, where hypometabolism significantly exceeded local levels of GMV or Aβ deposition. |
|  | Vannini et al. (2017) | Aβ-, n = 190 (72.8 ± 6.1) Aβ+, n = 61 (74.9 ± 6.2) | Greater SMCs correlated with decreased FDG metabolism in the bilateral precuneus, bilateral inferior parietal lobes, right inferior temporal lobe, right medial frontal gyrus, and right orbitofrontal gyrus. Decreased precuneus metabolism was associated with greater SMCs regardless of Aβ status, age, or thickness, whereas the relationship between hippocampal metabolism and SMCs was a function of Aβ, even after adjustment for age, hippocampal volume, or depressive symptoms. |
| Resting state fMRI^d^ | | | |
|  | Yang et al. (2019) | NC, n = 55 (63.41 ± 7.97) SCD, n = 43 (65.09 ± 8.66) aMCI, n = 52 (68.06 ± 9.32) d-AD, n = 44 (70.98 ± 10.02) | The significant main effects of frequency and group on fALFF differed widely across brain regions. The fALFF associated with primary disease effects was mainly distributed in the parietal lobe. Obvious frequency band and group interaction effects were observed in the left angular gyrus, left calcarine fissure and surrounding cortex, left superior cerebellum, left cuneus and right lingual gyrus. |
|  | Wang et al. (2019) | NC, n = 40 (68.07 ± 6.44) SCD, n = 32 (66.70 ± 5.98) aMCI, n = 37 (69.67 ± 7.48) AD, n = 30 (69.61 ± 9.53) | As compared to NC, SCD group showed decreased DC in the left somatomotor network and right frontoparietal control network. |
|  | Yang et al. (2018) | NC, n = 57 (63.77 ± 8.09) SCD, n = 44 (65.13 ± 8.57) aMCI, n = 55 (67.51 ± 9.62) d-AD, n = 47 (70.99 ± 10.07) | Compared with NC, SCD showed significantly decreased ALFF levels in the precuneus extend to PCC, hippocampus, cerebellum and lateral frontal cortex in the full band. |
|  | Li et al. (2018) | NC, n = 40 (75.10 ± 5.39) SMC, n = 44 (73.78 ± 5.81) | The voxel-wise comparison of Aβ deposition showed no significant difference between the SMC and NC groups. The SMC individuals showed higher DC in the bilateral hippocampus and left fusiform gyrus and lower DC in the inferior parietal region than NC. |
|  | Teipel et al. (2018) | NC, n = 80 (71.0 ± 4.2) SCD, n = 90 (72.0 ± 5.2) aMCI, n = 50 (72.5 ± 4.8) AD, n = 27 (72.7 ± 7.0) | The cross-validated discrimination accuracy for ALFF and functional connectivity ranged between 56% and 57% in the people with SCD. |
|  | Sun et al. (2016) | NC, n = 61 (64.11 ± 8.59) SCD, n = 25 (65.52 ± 6.12) | SCD exhibited higher ALFF values than NC in the bilateral inferior parietal lobule, right inferior and middle occipital gyrus, right superior temporal gyrus, and right cerebellum posterior lobe. |
| Tau-PET^e^ |  | | |
|  | Swinford et al. (2018) | NC, n = 40 (76.48 ± 7.21)  SMC, n = 11 (71.55 ± 5.11)  EMCI, n = 31 (75.32 ± 7.29) | In all participants, Self–ECog memory scores were significantly correlated with parahippocampal, frontal, parietal and global tau aggregation. |
|  | Buckley et al. (2017) | Clinically healthy participants, n = 133 (75.9± 7.0) | Greater SCD was associated with increasing entorhinal cortical tau burden, but not inferior temporal tau burden. After accounting for Aβ, the association between entorhinal cortical tau burden and SCD was largely unchanged. |

AD: Alzheimer disease; ALFF: amplitude of low frequency fluctuation; aMCI: amnestic mild cognitive impairment; ApoE: apolipoprotein; BPND: binding potential estimates relative to non-displaceable binding; CMRglc: cerebral metabolic rates for glucose; DC, degree centrality; ECog, Everyday cognition; EMCI, early mild cognitive impairment; FDG: ^18^F-fluorodeoxyglucose; GDS: Global global deterioration scale; GMV: gray matter volume; MCI: mild cognitive impairment; NC: normal control; ND: neurodegeneration; PIB: Pittsburgh compound B; ReHo: regional homogeneity; SCC: subjective cognitive concerns; SCD: subjective cognitive decline; SCI: subjective cognitive impairment; SMC: significant memory concerns; SMI: subjective memory impairment; SUVR: standardized uptake value ratio.

^a^To identify relevant amyloid PET studies, the PubMed were searched for studies published before October 2020 with search terms '((Subjective cognitive [Title/Abstract]) OR (Subjective memory [Title/Abstract]) OR (cognitive complaints [Title/Abstract])) AND (Alzheimer’s disease) AND (Pittsburgh or PiB or florbetapir or AV45 or florbetaben or flutemetamol or amyloid or abeta) ' and identified 322 studies. Studies focused on SCD in the context of AD and had voxel-wise or ROI based comparison of amyloid PET with NC would be included. 320 studies were excluded due to other topic or method based on review of title and abstract. Moreover, we add 2 studies manually.

^b^To identify relevant FDG-PET studies, the PubMed were searched for studies published before October 2020 with search terms ' ((Subjective cognitive [Title/Abstract]) OR (Subjective memory [Title/Abstract]) OR (cognitive complaints [Title/Abstract])) AND (Alzheimer’s disease) AND (Fluorodeoxyglucose OR FDG OR FDG-PET) ' and identified 55 studies. Studies focused on SCD in the context of AD and had voxel-wise or ROI based comparison of FDG PET with NC would be included. 50 studies were excluded due to other topic or method based on review of title and abstract. Moreover, we add 2 studies manually.

^c^To identify relevant amyloid and FDG PET studies, the PubMed were searched for studies published before October 2020 with search terms ‘((Subjective cognitive [Title/Abstract]) OR (Subjective memory [Title/Abstract]) OR (cognitive complaints [Title/Abstract])) AND (Alzheimer’s disease) AND (Pittsburgh or PiB or florbetapir or AV45 or florbetaben or flutemetamol or amyloid or abeta) AND (Fluorodeoxyglucose OR FDG OR FDG-PET)’ and identified 30 studies. Studies focused on SCD in the context of AD and had both amyloid and FDG PET would be included. 26 studies were excluded due to other topic or method based on review of title and abstract. Moreover, we add 1 study manually.

^d^To identify relevant fMRI studies, the PubMed were searched for studies published before October 2020 with search terms '((Subjective cognitive [Title/Abstract]) OR (Subjective memory [Title/Abstract]) OR (cognitive complaints [Title/Abstract])) AND (Alzheimer’s disease) AND (ALFF OR fALFF OR ReHo OR DC OR amplitude of low frequency fluctuation OR regional homogeneity OR degree centrality)' and identified 20 studies. Studies focused on SCD in the context of AD and had voxel-wise or ROI based comparison of ALFF, ReHo or DC with normal controls would be included. 14 studies were excluded due to other topic or method based on review of title and abstract.

^e^To identify relevant tau-PET studies, the PubMed were searched for studies published before October 2020 with search terms ' ((Subjective cognitive [Title/Abstract]) OR (Subjective memory [Title/Abstract]) OR (cognitive complaints [Title/Abstract])) AND (Alzheimer’s disease) AND (tau PET) ' and identified 33 studies. Studies focused on SCD in the context of AD would be included. 31 studies were excluded due to other topic or without tau-PET data based on review of title and abstract.

**Table S2. Partial correlation among neuroimaging metrics** **(i.e. AV45-SUVR, AV45-ALFF, AV45-ReHo, AV45-DC, FDG-SUVR, FDG-ALFF, FDG-ReHo, and FDG-DC) in the SCD group.**

|  |  | AV45-SUVR | AV45-ALFF | AV45-ReHo | AV45-  DC | FDG-SUVR | FDG-  ALFF | FDG-  ReHo | FDG-  DC |
| --- | --- | --- | --- | --- | --- | --- | --- | --- | --- |
| AV45-SUVR | *r*  [95% CI] | 1.00  [1.00, 1.00] |  |  |  |  |  |  |  |
|  | *P* | . |  |  |  |  |  |  |  |
| AV45-ALFF | *r*  [95% CI] | -0.18  [-0.42, 0.07] | 1.00  [1.00, 1.00] |  |  |  |  |  |  |
|  | *P* | 0.23 | . |  |  |  |  |  |  |
| AV45-ReHo | *r*  [95% CI] | -0.28  [-0.49, -0.04] | **0.75**  **[0.54, 0.86]** | 1.00  [1.00, 1.00] |  |  |  |  |  |
|  | *P* | 0.06 | **< 0.001** | . |  |  |  |  |  |
| AV45-DC | *r*  [95% CI] | -0.19  [-0.46, 0.13] | **0.70**  **[0.55, 0.83]** | **0.68**  **[0.49, 0.82]** | 1.00  [1.00, 1.00] |  |  |  |  |
|  | *P* | 0.20 | **< 0.001** | **< 0.001** | . |  |  |  |  |
| FDG-SUVR | *r*  [95% CI] | 0.20  [-0.13, 0.49] | -0.17  [-0.44, 0.20] | -0.11  [-0.38, 0.23] | -0.27  [-0.51, 0.08] | 1.00  [1.00, 1.00] |  |  |  |
|  | *P* | 0.21 | 0.28 | 0.48 | 0.09 | . |  |  |  |
| FDG-ALFF | *r*  [95% CI] | 0.07  [-0.26, 0.38] | **0.82**  **[0.57, 0.92]** | **0.62**  **[0.33, 0.79]** | **0.54**  **[0.26, 0.75]** | -0.20  [-0.54, 0.18] | 1.00  [1.00, 1.00] |  |  |
|  | *P* | 0.67 | **< 0.001** | **< 0.001** | **< 0.001** | 0.23 | . |  |  |
| FDG-ReHo | *r*  [95% CI] | -0.12  [-0.41, 0.24] | **0.56**  **[0.31, 0.73]** | **0.72**  **[0.56, 0.83]** | **0.66**  **[0.45, 0.80]** | -0.28  [-0.62, 0.14] | **0.63**  **[0.42, 0.78]** | 1.00  [1.00, 1.00] |  |
|  | *P* | 0.45 | **< 0.001** | **< 0.001** | **< 0.001** | 0.08 | **< 0.001** | . |  |
| FDG-DC | *r*  [95% CI] | -0.06  [-0.39, 0.32] | **0.45**  **[0.22, 0.68]** | **0.47**  **[0.28, 0.66]** | **0.63**  **[0.36, 0.80]** | -0.09  [-0.47, 0.31] | **0.55**  **[0.34, 0.76]** | **0.76**  **[0.60, 0.87]** | 1.00  [1.00, 1.00] |
|  | *P* | 0.74 | **0.005** | **0.003** | **< 0.001** | 0.59 | **< 0.001** | **< 0.001** | . |

Bold font indicated p < 0.05. AV45, [^18^F] florbetapir. FDG, [^18^F] fluorodeoxyglucose. SUVR, standardized uptake value ratio. ALFF, amplitude of low frequency fluctuation. ReHo, regional homogeneity. DC, degree centrality.CI, confidence interval.

**Table S3. Partial correlation between neuroimaging metrics (i.e. AV45-SUVR, AV45-ALFF, AV45-ReHo, AV45-DC, FDG-SUVR, FDG-ALFF, FDG-ReHo, and FDG-DC) and behavioral measures in the SCD group.**

|  |  | | AV45-SUVR | AV45-ALFF | AV45-ReHo | AV45-  DC | FDG-  SUVR | FDG-  ALFF | FDG-  ReHo | FDG-  DC |
| --- | --- | --- | --- | --- | --- | --- | --- | --- | --- | --- |
| AVLT-H delayed recall | | *r*  [95% CI] | -0.08  [-0.35, 0.19] | -0.10  [-0.44, 0.34] | 0.00  [-0.39, 0.40] | 0.06  [-0.29, 0.40] | 0.20  [-0.17, 0.48] | -0.08  [-0.42, 0.37] | 0.03  [-0.35, 0.39] | 0.30  [-0.04, 0.56] |
|  |  | *P* | 0.59 | 0.52 | 0.99 | 0.72 | 0.21 | 0.64 | 0.85 | 0.06 |
| AVLT-H recognition | | *r*  [95% CI] | 0.02  [-0.30, 0.33] | 0.04  [-0.21, 0.33] | 0.05  [-0.25, 0.34] | 0.14  [-0.13, 0.43] | 0.25  [-0.12, 0.51] | 0.09  [-0.25, 0.48] | 0.01  [-0.30, 0.36] | 0.27  [-0.02, 0.52] |
|  |  | *P* | 0.87 | 0.79 | 0.73 | 0.34 | 0.12 | 0.59 | 0.93 | 0.09 |
| STT part A | | *r*  [95% CI] | 0.11  [-0.17, 0.36] | 0.13  [-0.12, 0.36] | -0.02  [-0.23, 0.20] | 0.08  [-0.16, 0.29] | 0.04  [-0.35, 0.38] | 0.23  [-0.18, 0.56] | -0.19  [-0.46, 0.09] | -0.06  [-0.43, 0.23] |
|  |  | *P* | 0.46 | 0.39 | 0.91 | 0.60 | 0.79 | 0.15 | 0.25 | 0.72 |
| STT part B | | *r*  [95% CI] | **0.31**  **[0.30, 0.54]** | -0.05  [-0.30, 0.17] | -0.19  [-0.41, 0.03] | -0.18  [-0.43, 0.07] | 0.14  [-0.18, 0.42] | -0.02  [-0.37, 0.30] | -0.29  [-0.56, 0.08] | -0.12  [-0.45, 0.23] |
|  |  | *P* | **0.03** | 0.74 | 0.22 | 0.24 | 0.36 | 0.91 | 0.08 | 0.48 |
| AFT | | *r*  [95% CI] | -0.10  [-0.33, 0.16] | -0.02  [-0.30, 0.26] | 0.05  [-0.24, 0.32] | 0.09  [-0.19, 0.35] | -0.17  [-0.46, 0.15] | 0.05  [-0.23, 0.33] | -0.08  [-0.38, 0.20] | -0.02  [-0.36, 0.28] |
|  |  | *P* | 0.49 | 0.89 | 0.76 | 0.55 | 0.29 | 0.78 | 0.63 | 0.92 |
| BNT | | *r*  [95% CI] | -0.04  [-0.34, 0.24] | -0.26  [-0.48, 0.00] | -0.07  [-0.34, 0.22] | -0.20  [-0.46, 0.08] | 0.07  [-0.30, 0.41] | **-0.32**  **[-0.60, 0.12]** | -0.06  [-0.36, 0.32] | -0.16  [-0.51, 0.31] |
|  |  | *P* | 0.78 | 0.08 | 0.65 | 0.18 | 0.65 | **0.046** | 0.74 | 0.34 |
| MoCA-B | | *r*  [95% CI] | -0.12  [-0.41, 0.15] | -0.11  [-0.36, 0.14] | 0.17  [-0.12, 0.43] | 0.14  [-0.12, 0.39] | -0.23  [-0.47, 0.02] | 0.05  [-0.23, 0.40] | 0.23  [-0.09, 0.50] | 0.20  [-0.11, 0.54] |
|  |  | *P* | 0.43 | 0.47 | 0.25 | 0.36 | 0.15 | 0.77 | 0.16 | 0.23 |
| HAMD | | *r*  [95% CI] | 0.09  [-0.16, 0.39] | -0.04  [-0.26, 0.22] | -0.07  [-0.26, 0.13] | -0.14  [-0.39, 0.18] | -0.02  [-0.26, 0.22] | 0.25  [0.03, 0.52] | 0.09  [-0.13, 0.35] | 0.15  [-0.13, 0.44] |
|  |  | *P* | 0.53 | 0.80 | 0.63 | 0.35 | 0.92 | 0.13 | 0.59 | 0.37 |
| HAMA | | *r*  [95% CI] | 0.03  [-0.19, 0.29] | -0.26  [-0.46, 0.01] | -0.19  [-0.36, 0.03] | -0.25  [-0.50, 0.07] | 0.21  [-0.06, 0.47] | -0.03  [-0.30, 0.23] | 0.02  [-0.26, 0.29] | 0.07  [-0.29, 0.39] |
|  |  | *P* | 0.85 | 0.09 | 0.21 | 0.09 | 0.17 | 0.84 | 0.91 | 0.66 |

Bold font indicated p < 0.05. AVLT-H, auditory verbal learning test-Huashan version. STT, shape trails test. AFT, animal fluency test. BNT, Boston naming test. MoCA-B, Montreal cognitive assessment-basic. HAMD, Hamilton depression scale. HAMA, Hamilton anxiety scale. AV45, [^18^F] florbetapir. FDG, [^18^F] fluorodeoxyglucose. SUVR, standardized uptake value ratio. ALFF, amplitude of low frequency fluctuation. ReHo, regional homogeneity. DC, degree centrality. CI, confidence interval.

**Table S4. Demographies and neuropsychological tests of the SCD^+^, SCD^-^ and NC^-^ groups.**

|  | SCD^+^ (N = 28) | SCD^-^ (N = 22) | NC^-^ (N = 11) | Statistics |
| --- | --- | --- | --- | --- |
| Age (years) | 66.9 ± 6.2 | 65.8 ± 5.2 | 67.5 ± 4.4 | *F_(2,58)_* = 0.45, *P* = 0.64 |
| Gender (M/F) | 7/21 | 12/10 | 3/8 | *χ^2^* = 5.11, *P* = 0.08 |
| Education (years) | 13.3 ± 3.0 | 14.4 ± 2.9 | 12.7 ± 2.6 | *F_(2,58)_* = 1.41, *P* = 0.25 |
| HAMD | 4.0 ± 3.8 | 3.6 ± 3.8 | 0.8 ± 1.2 | *F_(2,58)_* = 3.53, *P* = 0.04^bc^ |
| HAMA | 4.5 ± 3.0 | 4.4 ± 4.9 | 1.5 ± 2.4 | *F_(2,58)_* = 2.78, *P* = 0.07^bc^ |
| AVLT-H delayed recall | 8.0 ± 1.8 | 7.4 ± 2.2 | 7.2 ± 2.0 | *F_(2,58)_* = 1.06, *P* = 0.35 |
| AVLT-H recognition | 22.3 ± 1.2 | 22.4 ± 1.6 | 22.3 ± 1.7 | *F_(2,58)_* = 0.05, *P* = 0.95 |
| STT part A | 55.8 ± 20.8 | 55.2 ± 13.4 | 53.3 ± 14.8 | *F_(2,58)_* = 0.08, *P* = 0.92 |
| STT part B | 134.6 ± 41.8 | 121.2 ± 29.2 | 132.7 ± 28.4 | *F_(2,58)_* = 0.94, *P* = 0.40 |
| AFT | 20.7 ± 5.2 | 19.1 ± 3.9 | 19.1 ± 5.2 | *F_(2,58)_* = 0.84, *P* = 0.44 |
| BNT | 25.8 ± 2.6 | 26.3 ± 2.1 | 25.0 ± 3.7 | *F_(2,58)_* = 0.88, *P* = 0.42 |
| MoCA-B | 26.5 ± 2.2 | 26.1 ± 2.0 | 25.6 ± 2.0 | *F_(2,58)_* = 0.80, *P* = 0.45 |

Data were presented as the mean ± SD. SCD, subjective cognitive decline. NC, normal control. SCD^+^, Aβ positive SCD. SCD^-^, Aβ negative SCD. NC^-^, Aβ negative NC. HAMD, Hamilton depression scale. HAMA, Hamilton anxiety scale. AVLT-H, auditory verbal learning test-Huashan version. STT, shape trails test. AFT, animal fluency test. BNT, Boston naming test. MoCA-B, Montreal cognitive assessment-basic.

^a^Indicates significant differences between the SCD^+^ and SCD^-^ group; ^b^Indicates significant differences between the SCD^+^ and NC^-^ group; ^c^Indicates significant differences between the SCD^-^ and NC^-^ group.

**Table S5. Partial correlation between neuroimaging metrics (i.e. AV45-SUVR, AV45-ALFF, AV45-ReHo, AV45-DC, FDG-SUVR, FDG-ALFF, FDG-ReHo, and FDG-DC) in the SCD group from independent verification of data analyses.**

|  |  | AV45-SUVR | AV45-ALFF | AV45-ReHo | AV45-DC | FDG-SUVR | FDG-ALFF | FDG-ReHo | FDG-DC |
| --- | --- | --- | --- | --- | --- | --- | --- | --- | --- |
| AV45-SUVR | *r*  [95% CI] | 1.00  [1.00, 1.00] |  |  |  |  |  |  |  |
|  | *P* | . |  |  |  |  |  |  |  |
| AV45-ALFF | *r*  [95% CI] | -0.19  [-0.43, 0.08] | 1.00  [1.00, 1.00] |  |  |  |  |  |  |
|  | *P* | 0.21 | . |  |  |  |  |  |  |
| AV45-ReHo | *r*  [95% CI] | -0.31  [-0.51, -0.04] | **0.74**  **[0.55, 0.86]** | 1.00  [1.00, 1.00] |  |  |  |  |  |
|  | *P* | 0.04 | **< 0.001** | . |  |  |  |  |  |
| AV45-DC | *r*  [95% CI] | -0.22  [-0.47, 0.09] | **0.70**  **[0.54, 0.83]** | **0.66**  **[0.45, 0.81]** | 1.00  [1.00, 1.00] |  |  |  |  |
|  | *P* | 0.15 | **< 0.001** | **< 0.001** | . |  |  |  |  |
| FDG-SUVR | *r*  [95% CI] | 0.20  [-0.11, 0.48] | -0.20  [-0.48, 0.18] | -0.10  [-0.39, 0.22] | -0.29  [-0.52, 0.03] | 1.00  [1.00, 1.00] |  |  |  |
|  | *P* | 0.21 | 0.20 | 0.53 | 0.07 | . |  |  |  |
| FDG-ALFF | *r*  [95% CI] | 0.06  [-0.24, 0.43] | **0.80**  **[0.54, 0.91]** | **0.54**  **[0.24, 0.74]** | **0.49**  **[0.20, 0.72]** | -0.21  [-0.53, 0.21] | 1.00  [1.00, 1.00] |  |  |
|  | *P* | 0.70 | **< 0.001** | **< 0.001** | **0.002** | 0.19 | . |  |  |
| FDG-ReHo | *r*  [95% CI] | -0.13  [-0.43, 0.25] | **0.54**  **[0.31, 0.70]** | **0.68**  **[0.53, 0.81]** | **0.61**  **[0.37, 0.78]** | -0.28  [-0.61, 0.15] | **0.56**  **[0.32, 0.75]** | 1.00  [1.00, 1.00] |  |
|  | *P* | 0.42 | **< 0.001** | **< 0.001** | **< 0.001** | 0.08 | **< 0.001** | . |  |
| FDG-DC | *r*  [95% CI] | -0.07  [-0.39, 0.33] | **0.41**  **[0.13, 0.64]** | **0.43**  **[0.19, 0.62]** | **0.59**  **[0.31, 0.78]** | -0.10  [-0.48, 0.29] | **0.49**  **[0.27, 0.69]** | **0.74**  **[0.55, 0.86]** | 1.00  [1.00, 1.00] |
|  | *P* | 0.68 | **0.01** | **0.008** | **< 0.001** | 0.56 | **0.002** | **< 0.001** | . |

The dPCu-ROI used in the independent verification of data analyses (by authors CC Ding and JH Jiang) were same with the ROI shown in the Figure 1 (6 voxels). Notably, the metric values extracted from the ROI were derived from the independent verification results. Bold font indicated p < 0.05. AV45, [^18^F] florbetapir. FDG, [^18^F] fluorodeoxyglucose. SUVR, standardized uptake value ratio. ALFF, amplitude of low frequency fluctuation. ReHo, regional homogeneity. DC, degree centrality. CI, confidence interval.

**Table S6. Partial correlation between** **neuroimaging metrics (i.e. AV45-SUVR, AV45-ALFF, AV45-ReHo, AV45-DC, FDG-SUVR, FDG-ALFF, FDG-ReHo, and FDG-DC) and behavioral measures in the SCD group** **from independent verification of data analyses.**

|  |  | AV45-SUVR | AV45-ALFF | AV45-ReHo | | AV45-DC | | FDG-SUVR | | FDG-ALFF | | FDG-ReHo | | FDG-DC |
| --- | --- | --- | --- | --- | --- | --- | --- | --- | --- | --- | --- | --- | --- | --- |
| AVLT-H delayed recall | *r*  [95% CI] | -0.08  [-0.36, 0.19] | -0.10  [-0.45, 0.32] | 0.03  [-0.35, 043] | | 0.09  [-0.26, 0.42] | | 0.20  [-0.17, 0.49] | | -0.06  [-0.38, 0.38] | | 0.05  [-0.30, 0.39] | | **0.34**  **[0.01, 0.60]** |
|  | *P* | 0.59 | 0.52 | 0.86 | | 0.55 | | 0.21 | | 0.70 | | 0.78 | | **0.04** |
| AVLT-H recognition | *r*  [95% CI] | 0.03  [-0.28, 0.32] | 0.05  [-0.22, 0.34] | 0.08  [-0.21, 0.36] | | 0.19  [-0.13, 0.46] | | 0.25  [-0.14, 0.52] | | 0.09  [-0.25, 0.48] | | 0.01  [-0.28, 0.33] | | 0.29  [0.03, 0.55] |
|  | *P* | 0.87 | 0.76 | 0.61 | | 0.22 | | 0.12 | | 0.58 | | 0.98 | | 0.07 |
| STT part A | *r*  [95% CI] | 0.11  [-0.18, 0.36] | 0.13  [-0.14, 0.37] | -0.01  [-0.22, 0.19] | | 0.09  [-0.15, 0.30] | | 0.04  [-0.33, 0.37] | | 0.25  [-0.18, 0.59] | | -0.24  [-0.53, 0.07] | | -0.09  [-0.42, 0.19] |
|  | *P* | 0.45 | 0.39 | 0.95 | | 0.54 | | 0.80 | | 0.13 | | 0.14 | | 0.59 |
| STT part B | *r*  [95% CI] | **0.31**  **[0.05, 0.54]** | -0.05  [-0.30, 0.20] | -0.16  [-0.41, 0.08] | | -0.15  [-0.41, 0.10] | | 0.14  [-0.16, 0.41] | | 0.01  [-0.33, 0.34] | | -0.29  [-0.59, 0.06] | | -0.08  [-0.37, 0.25] |
|  | *P* | **0.03** | 0.73 | 0.28 | | 0.31 | | 0.37 | | 0.95 | | 0.08 | | 0.64 |
| AFT | *r*  [95% CI] | -0.10  [-0.35, 0.17] | -0.26  [-0.48, 0.03] | 0.02  [-0.28, 0.28] | | 0.11  [-0.19, 0.34] | | -0.17  [-0.46, 0.11] | | 0.10  [-0.20, 0.37] | | -0.10  [-0.39, 0.16] | | 0.04  [-0.28, 0.35] |
|  | *P* | 0.49 | 0.08 | 0.91 | | 0.47 | | 0.29 | | 0.56 | | 0.56 | | 0.83 |
| BNT | *r*  [95% CI] | -0.04  [-0.36, 0.22] | -0.26  [-0.48, 0.03] | -0.07  [-0.32, 0.18] | | -0.17  [-0.42, 0.10] | | 0.07  [-0.29, 0.39] | | **-0.32**  **[-0.59, 0.06]** | | -0.03  [-0.36, 0.36] | | -0.14  [-0.47, 0.31] |
|  | *P* | 0.78 | 0.08 | 0.65 | | 0.26 | | 0.64 | | **0.047** | | 0.87 | | 0.40 |
| MoCA-B | *r*  [95% CI] | -0.12  [-0.45, 0.22] | -0.10  [-0.36, 0.12] | 0.14  [-0.12, 0.39] | | 0.14  [-0.13, 0.40] | | -0.23  [-0.46, 0.04] | | 0.02  [-0.27, 0.34] | | 0.18  [-0.12, 0.48] | | 0.18  [-0.14, 0.51] |
|  | *P* | 0.19 | 0.50 | 0.35 | | 0.34 | | 0.15 | | 0.91 | | 0.26 | | 0.27 |
| HAMD | *r*  [95% CI] | 0.09  [-0.15, 0.39] | -0.07  [-0.27, 0.19] | -0.06  [-0.26, 0.19] | | -0.16  [-0.41, 0.14] | | -0.02  [-0.25, 0.22] | | 0.23  [0.00, 0.51] | | 0.08  [-0.15, 0.38] | | 0.15  [-0.08, 0.47] |
|  | *P* | 0.53 | 0.66 | 0.69 | | 0.29 | | 0.92 | | 0.16 | | 0.65 | | 0.37 |
| HAMA | *r*  [95% CI] | 0.03  [-0.18, 0.29] | **-0.29**  **[-0.49, -0.06]** | -0.21  [-0.38, 0.01] | -0.29  [-0.51, 0.07] | | 0.22  [-0.05, 0.44] | | -0.07  [-0.31, 0.18] | | -0.03  [-0.30, 0.24] | | 0.07  [-0.28, 0.39] | |
|  | *P* | 0.85 | **0.048** | 0.17 | 0.05 | | 0.17 | | 0.69 | | 0.88 | | 0.66 | |

The dPCu-ROI used in the independent verification of data analyses (by authors CC Ding and JH Jiang) were same with the ROI shown in the Figure 1 (6 voxels). Notably, the metric values extracted from the ROI were derived from the independent verification results. Bold font indicated p < 0.05. AVLT-H, auditory verbal learning test-Huashan version. STT, shape trails test. AFT, animal fluency test. BNT, Boston naming test. MoCA-B, Montreal cognitive assessment-basic. HAMD, Hamilton depression scale. HAMA, Hamilton anxiety scale. AV45, [^18^F] florbetapir. FDG, [^18^F] fluorodeoxyglucose. SUVR, standardized uptake value ratio. ALFF, amplitude of low frequency fluctuation. ReHo, regional homogeneity. DC, degree centrality. CI, confidence interval.

**Table S7. Summary of SCD-related task fMRI studies.**

| References | Task | Results |
| --- | --- | --- |
| Rodda et al., 2009 | Verbal episodic memory encoding task | The SCD group showed greater activation in the left prefrontal cortex during memory task. |
| Dumas et al., 2013 | Visual-verbal N-back test | Compared with the NC group, the SCD group showed greater activation in the middle frontal gyrus, the precuneus and the cingulate gyrus. |
| Rodda et al., 2011 | Divided attention task | The SCD group showed greater activation in left medial temporal lobe, bilateral thalamus, posterior cingulate and caudate during the divided attention task. |
| Erk et al., 2011 | Episodic Memory Task  N-Back Task | The SCD group was associated with a reduction in right hippocampal activation and increased activation in the right dorsolateral prefrontal cortex during episodic memory recall task. No difference in brain activation during working memory were observed in the SCD group. |
| Hayes et al., 2017 | Event-related visual memory encoding task. | SCD showed lower subsequent memory effects in the occipital lobe, superior parietal lobe, and posterior cingulate cortex. |
| Hu et al., 2017 | Intertemporal decision task, with and without simultaneous episodic future imagination | SCD showed reduced future-oriented choices and related to reduced hippocampal activation. |

**Table S8. Summary of studies of correlation analyses of PET glucose metabolism and RS-fMRI local metrics (ReHo, ALFF, DC) in healthy subjects.**

| References | Correlation across-voxels* | Correlation across-subjects** |
| --- | --- | --- |
| Aiello et al., 2015 | Very significant | Only a few regions  Only a few regions  Only a few regions  Almost no |
| Bernier M et al., 2017 in healthy young subjects | Very significant |  |
| Bernier M et al., 2017 in healthy aged subjects | Very significant |  |
| Jiao F et al., et al., 2019 | Very significant |  |

*: Correlation is performed across-voxels for each individual. The *N* is the number of voxels in the brain.

**: Correlation is performed across-subjects for each voxel. The *N* is the number of subjects.

**Table S9. Summary of studies reporting abnormal PET glucose metabolism and RS-fMRI metrics in brain disorders.**

| Region in a disorder | Glucose or  RS-fMRI | Increase(↑) or decrease(↓) | References |
| --- | --- | --- | --- |
| Epileptic focus of the temporal lobe epilepsy | Glucose | ↓ | Guedj E et al., 2015 |
|  | ALFF | ↑ | Zhang ZQ et al., 2010; Ji GJ et al., 2013 |
| Putamen in Parkinson’s disease | Glucose | ↑ | Tomše P et al., 2017 |
|  | ALFF | ↓ | Wang J et al., 2018 |
| PCC in AD/MCI | Glucose | ↓ | He W et al., 2015 |
|  | ALFF | ↓ | Pan et al., 2016 |
| Dorsal PCu in SCD | Glucose | ↑ | The current study |
|  | ALFF, ReHo, DC | ↓ | The current study |

**Table S10. The subject numbers included in the statistical analysis for each modality, i.e. AV45-PET, FDG-PET, AV45-fMRI, and FDG-fMRI.**

|  |  | SCD (N) | NC (N) | Reasons for exclusion |
| --- | --- | --- | --- | --- |
| AV45-PET | raw data | 50 | 15 |  |
|  | excluded | 0 | 0 |  |
| FDG-PET | raw data | 45 | 14 | One NC and five SCD subjects lacked raw FDG-PET data. |
|  | excluded | 0 | 0 |  |
| AV45-fMRI | raw data | 49 | 13 | One SCD subject was excluded because of large head motion (more than 3 mm of translation or 3° of rotation in any direction). Two NC subjects were excluded due to substantial distortion (Fig. S8). |
|  | excluded | 1 | 2 |  |
| FDG-fMRI | raw data | 42 | 14 | One NC and five SCD subjects lacked raw FDG-fMRI data. Three SCD subjects were excluded because of large head motion. |
|  | excluded | 3 | 0 |  |

SCD, subjective cognitive decline. NC, normal control. PET, positron emission tomography. AV45, [^18^F] florbetapir. FDG, [^18^F] fluorodeoxyglucose. fMRI, functional magnetic resonance imaging.

**Supplementary Figures**

**Figure S1.** **Eligibility criteria for the study population**

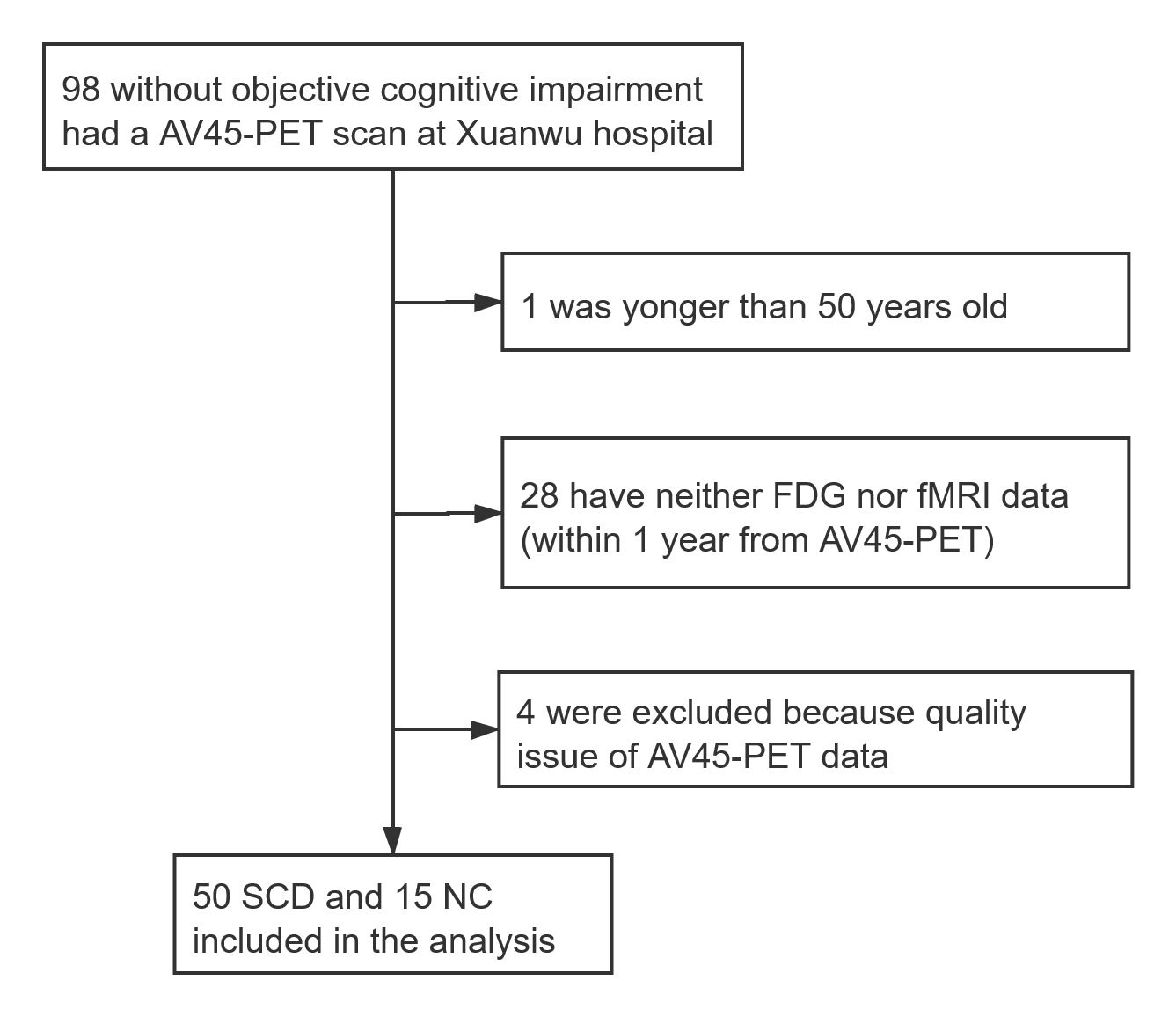

**Figure S2. Group differences between SCD and NC groups with controlling for age, sex, and years of education, on Aβ deposition and glucose metabolism (top row) and RS-fMRI metrics (bottom row) from independent verification of data analyses (by authors CC Ding and JH Jiang).**

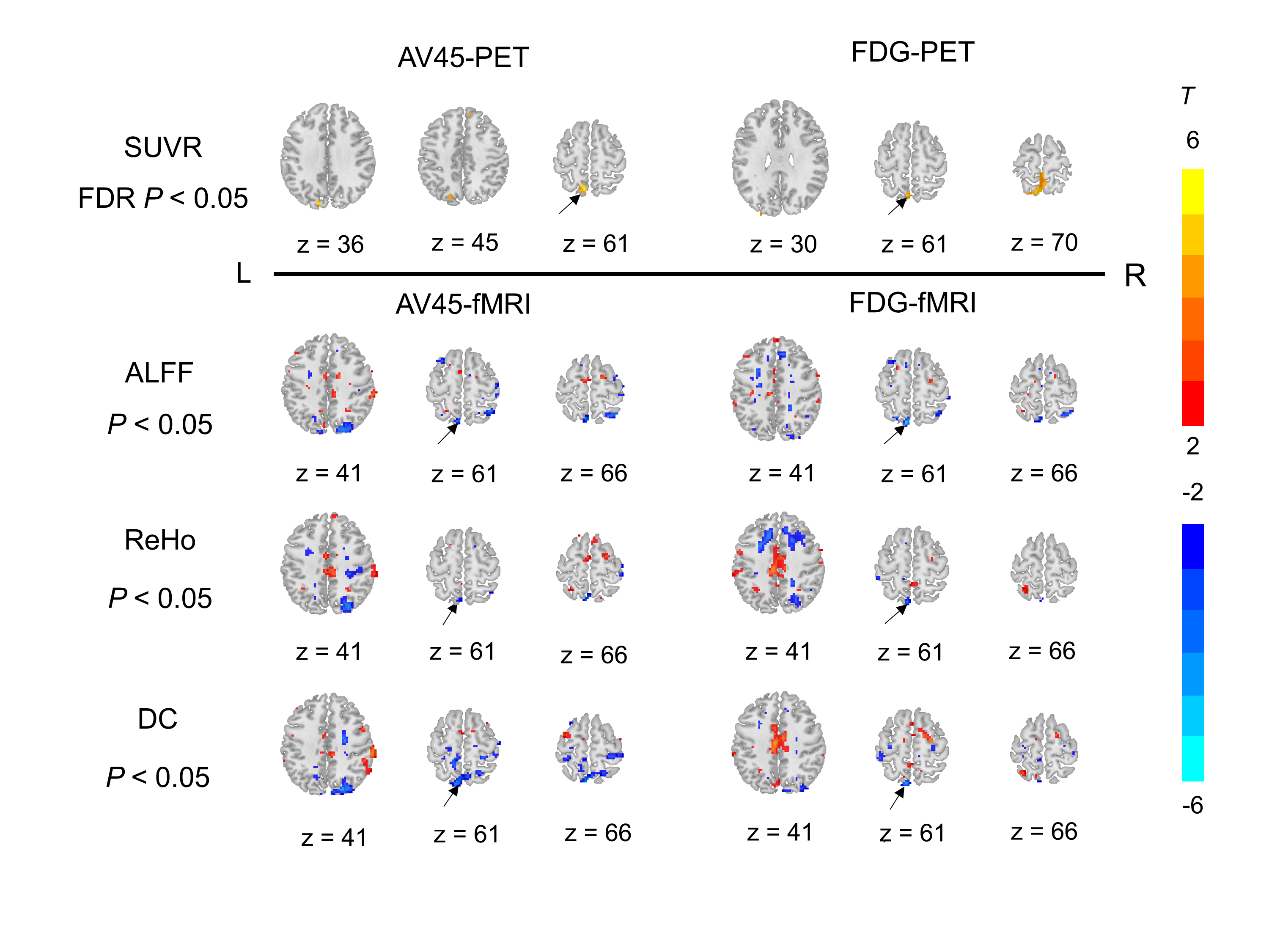

Red and yellow colors indicate increased metrics in SCD vs. NC, and blue color indicates the opposite. SCD, subjective cognitive decline. NC, normal control. dPCu, dorsal PCu. AV45, [^18^F] florbetapir. FDG, [^18^F] fluorodeoxyglucose. SUVR, standardized uptake value ratio. FDR, false discovery rate. ALFF, amplitude of low frequency fluctuation. ReHo, regional homogeneity. DC, degree centrality. L: left in the brain. R: right in the brain. *T*: the *T* value of two-sample *t*-test on the PET and fMRI metrics maps.

**Figure S3.** **Patterns of seed-based resting-state functional connectivity (RSFC) of the left dPCu of all the subjects in AV45-fMRI dataset (A) and FDG-fMRI dataset (B) (*P* < 0.05, FDR corrected) from independent verification of data analyses (by authors CC Ding and JH Jiang).**

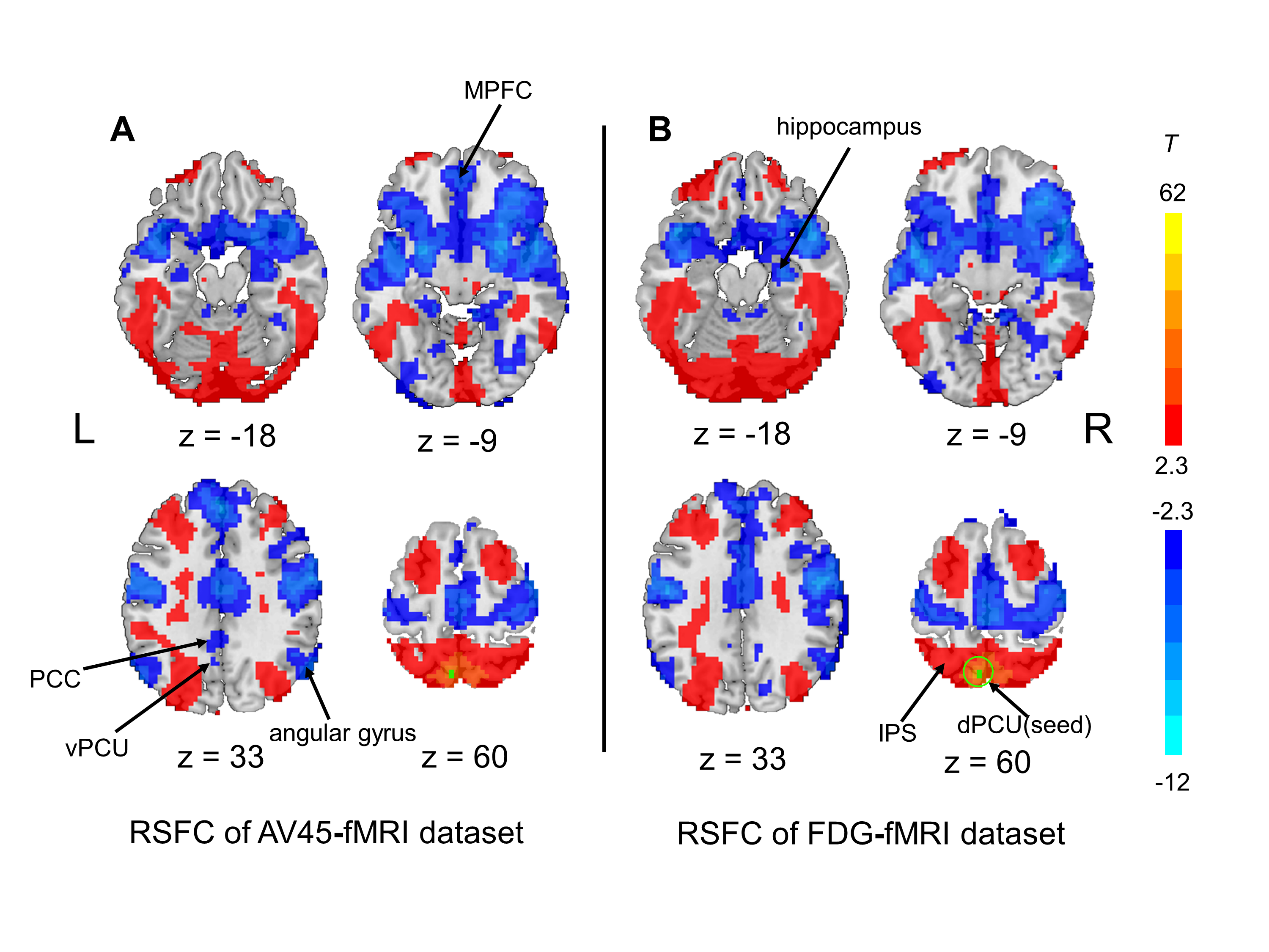

The yellow-red color indicates positive connections and the blue color indicates that negative connections with the left dorsal PCu. MPFC, medial prefrontal cortex. PCC, posterior cingulate cortex. vPCu, ventral PCu. dPCu, dorsal PCu. IPS, intraparietal sulcus. L: left in the brain. R: right in the brain. *T*: the *T* value of one-sample *t*-test on the RSFC maps.

**Figure S4.** **Comparisons of global AV45-SUVR and the 8 metrics in the dPCu-ROI among three groups from independent verification of data analyses, i.e. AV45-SUVR (B), FDG-SUVR (C), AV45-ALFF (D), AV45-ReHo (E), AV45-DC (F), FDG-ALFF (G), FDG-ReHo (H), and FDG-DC (I) (by authors CC Ding and JH Jiang).**

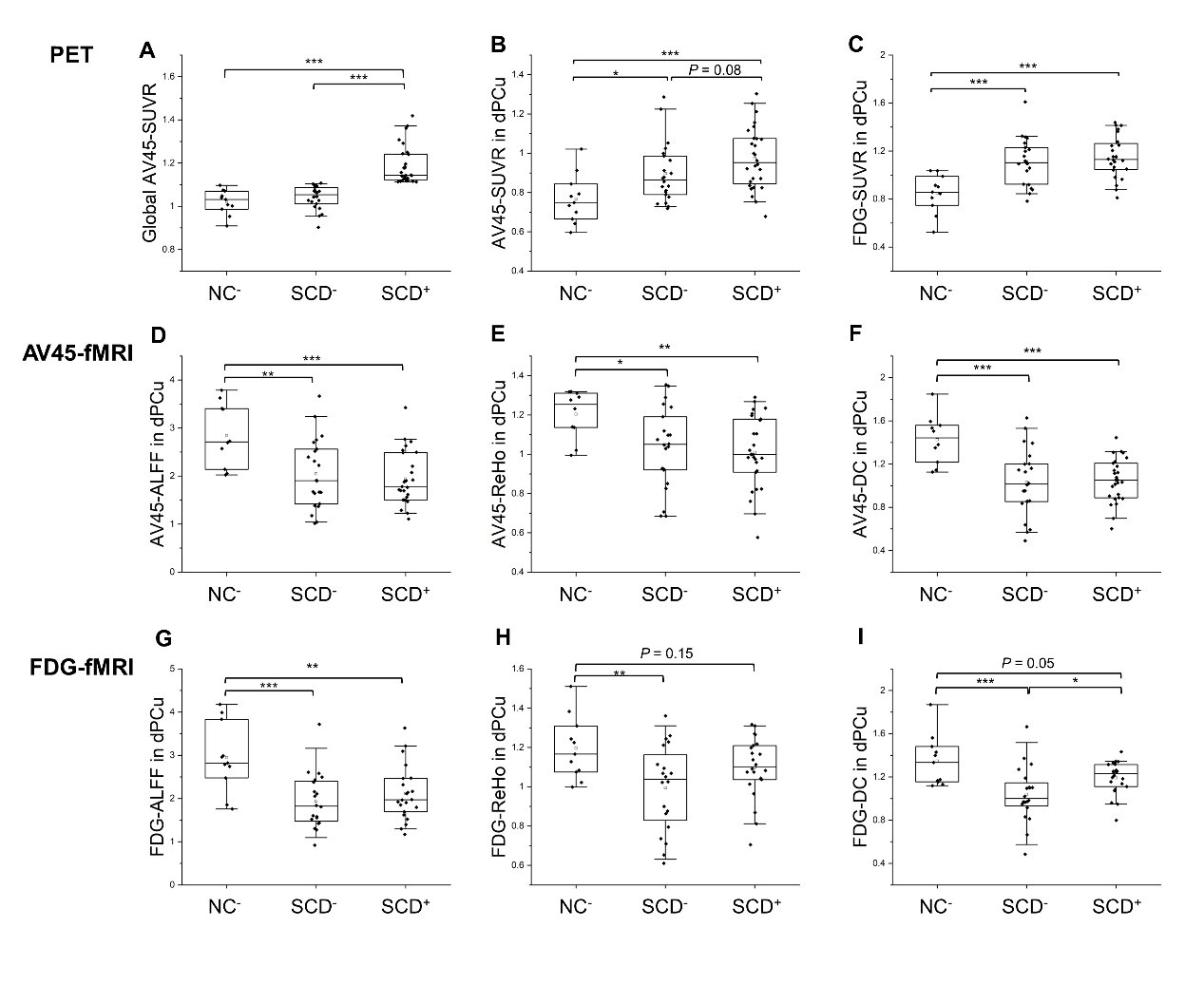

The dPCu-ROI used in the independent verification of data analyses were same with the ROI shown in the Figure 1 (6 voxels). NC^-^, Aβ negative NC. SCD^-^, Aβ negative SCD. SCD^+^, Aβ positive SCD. *: *P* < 0.05, **: *P* < 0.01, ***: *P* < 0.001. All box and whisker plots: box range, 25–75%; whisker range, 5–95%.

**Figure S5.** **Simulation of the correlation between mean value and std value.**

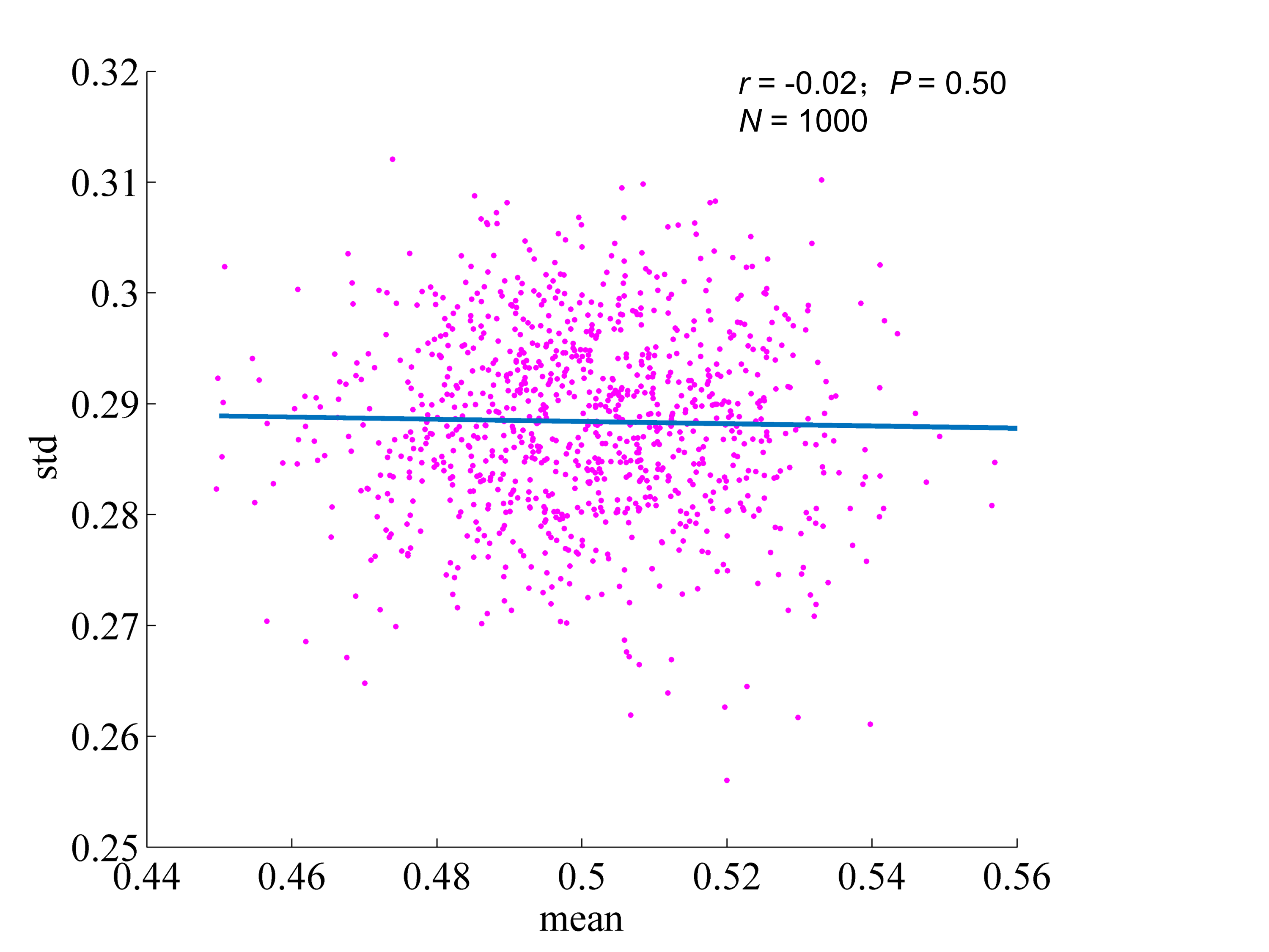

The simulation was done as follows: steps. 1000 signal courses with 240 time points containing pseudorandom values were generated from the standard uniform distribution on the open interval (0,1), and then the mean and standard deviation values were computed.

**Figure S6. Comparisons of the 8 metrics in the PCC-ROI (A) among three groups, i.e. AV45-SUVR (B), FDG-SUVR (C), AV45-ALFF (D), AV45-ReHo (E), AV45-DC (F), FDG-ALFF (G), FDG-ReHo (H), and FDG-DC (I).**

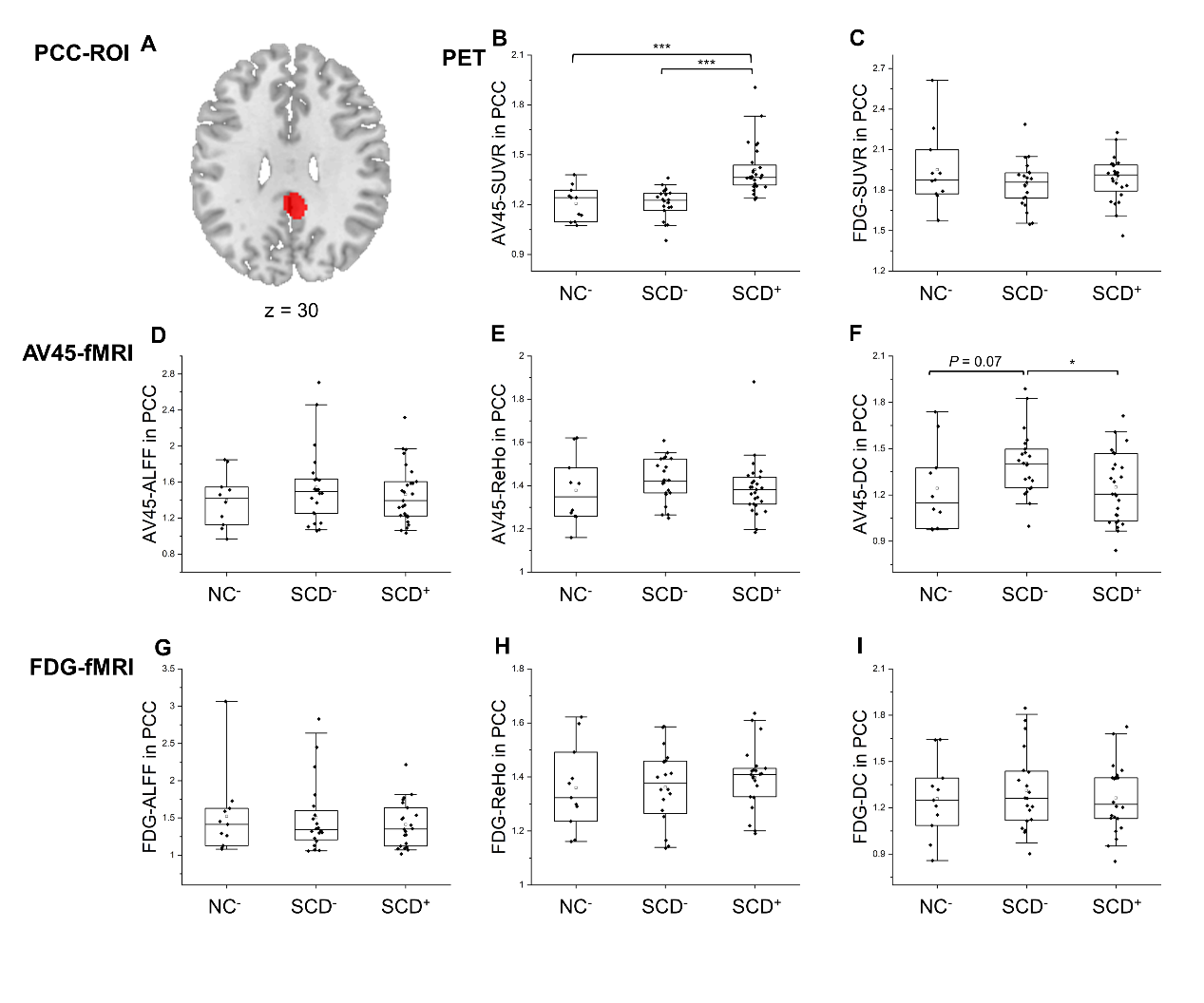

NC^-^, Aβ negative NC. SCD^-^, Aβ negative SCD. SCD^+^, Aβ positive SCD. *: *P* < 0.05, **: *P* < 0.01, ***: *P* < 0.001. All box and whisker plots: box range, 25–75%; whisker range, 5–95%.

**Figure S7. AV45-PET images with quality issue**

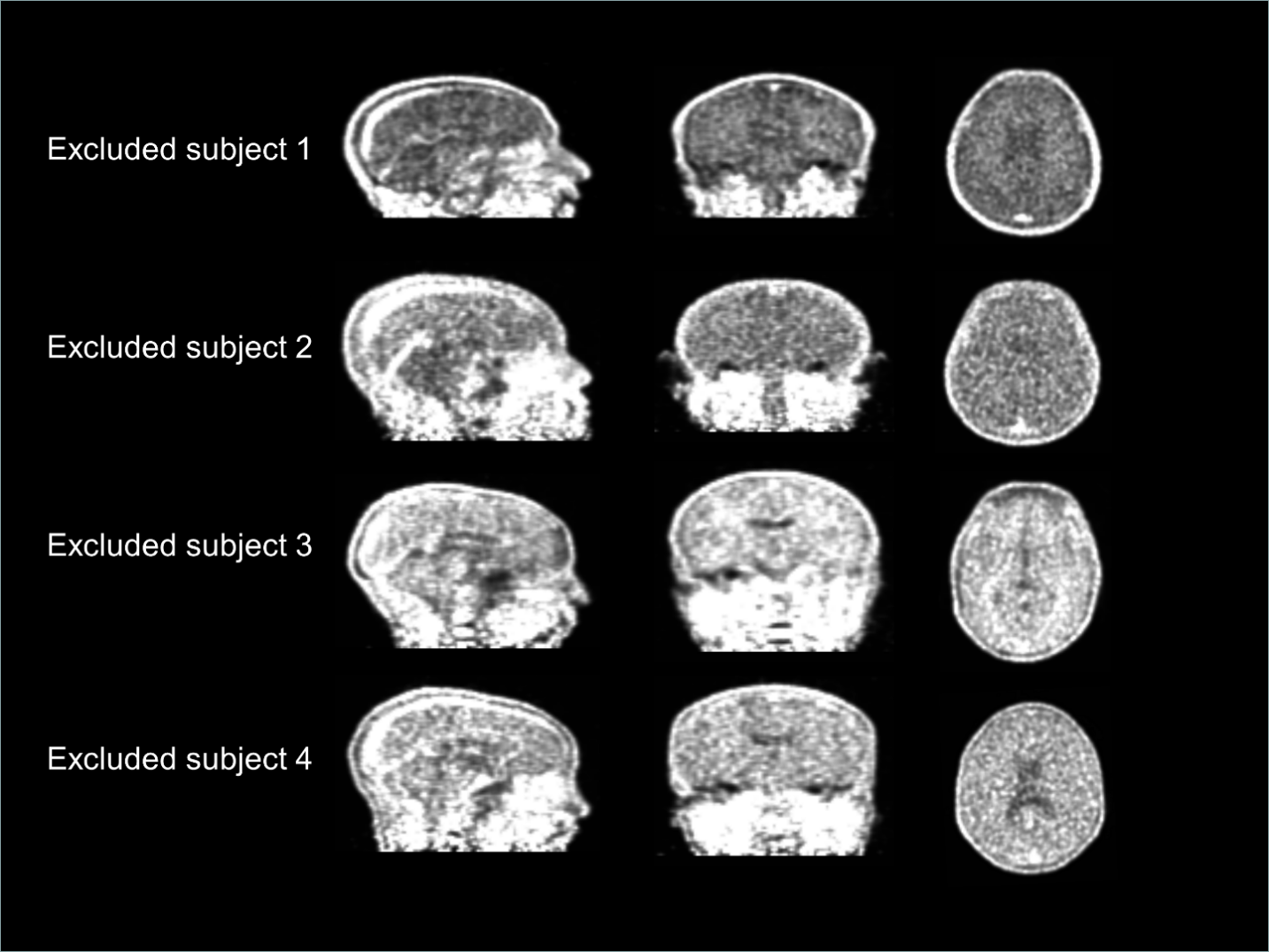

Four SCD subjects were excluded from this study due to quality issue of their AV45-PET images. That is, a total of 65 subjects (50 for SCD and 15 for NC) were included. AV45, [^18^F] florbetapir. SCD, subjective cognitive decline. NC, normal control.

**Figure S8.** **AV45-fMRI images with substantial signal loss and distortion.**

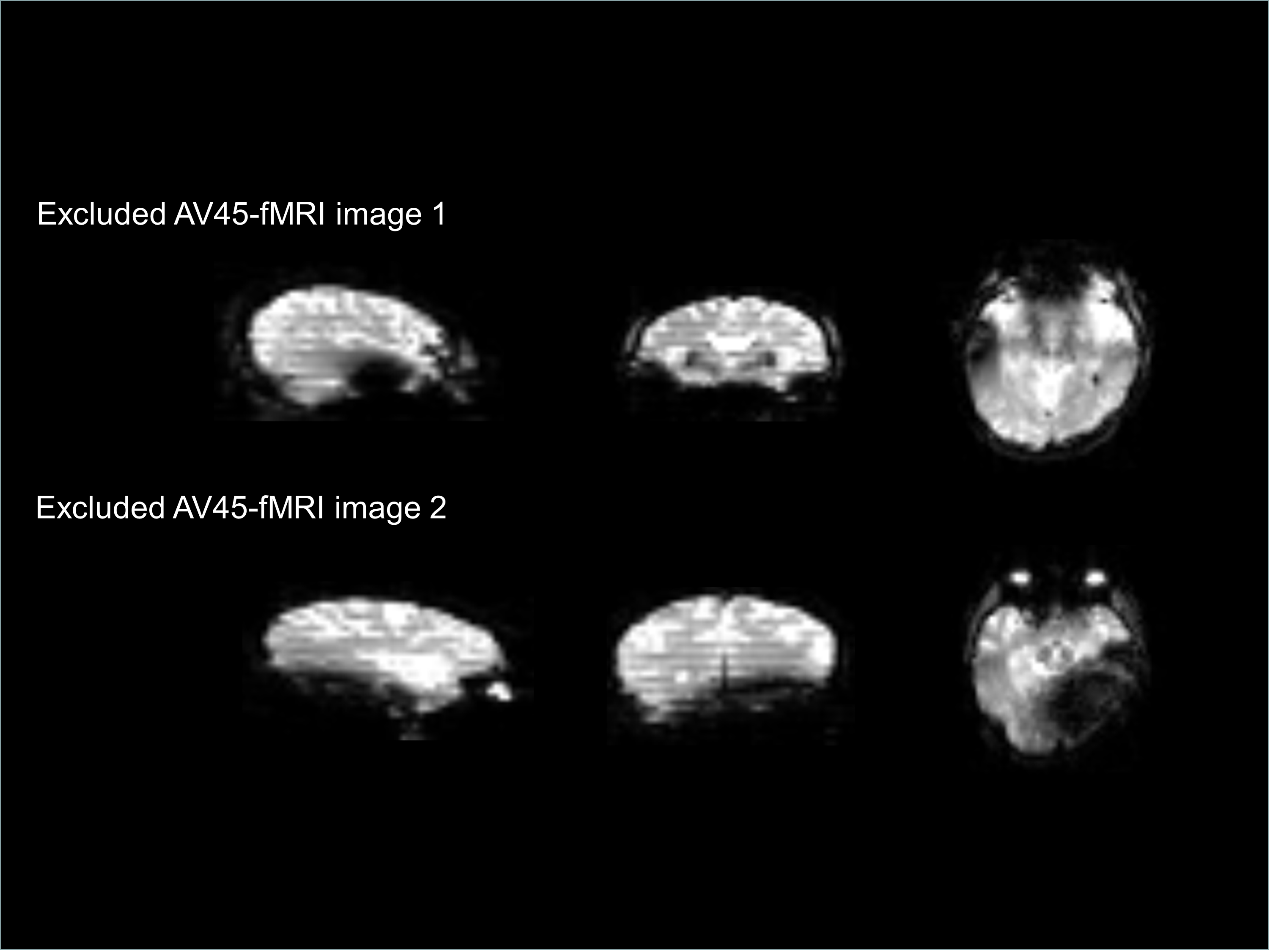

**Supplementary Appendix References**

1. Ashburner J, Friston KJ. Unified segmentation. Neuroimage 2005;26:839–851.
2. Landau SM, Mintun MA, Joshi AD, et al. Alzheimer's Disease Neuroimaging Initiative. Amyloid deposition, hypometabolism, and longitudinal cognitive decline. Ann Neurol 2012;72:578-586.
3. Landau SM, Breault C, Joshi AD, et al. Alzheimer’s Disease Neuroimaging Initiative. Amyloid-β imaging with Pittsburgh compound B and florbetapir: comparing radiotracers and quantification methods. J Nucl Med 2013;54:70-77.
4. Yan CG, Wang XD, Zuo XN, Zang YF. DPABI: Data Processing & Analysis for (Resting-State) Brain Imaging. Neuroinformatics 2016;14:339–351.
5. Jia XZ, Wang J, Sun HY, et al. RESTplus:an improved toolkit for resting-state functional magnetic resonance imaging data processing. Sci Bull 2019;64:953-954.
6. Zang YF, He Y, Zhu CZ, et al. Altered baseline brain activity in children with ADHD revealed by resting-state functional MRI. Brain Dev 2007;29:83-91.
7. Zang YF, Jiang T, Lu Y, He Y, Tian L. Regional homogeneity approach to fMRI data analysis. Neuroimage 2004;22:394-400.
8. Zuo XN, Ehmke R, Mennes M, et al. Network centrality in the human functional connectome. Cereb Cortex 2012;22:1862-1875.
